# Epidemiological Risk Factors of SARS-CoV-2 Infections

**DOI:** 10.1101/2020.09.30.20204990

**Authors:** William A. Barletta

## Abstract

Since the first recognitions by governments of the pandemic characteristic of the SARS-CoV-2 infections, public health agencies have warned the public about the dangers of the virus to persons with a variety of underlying physical conditions, many of which are more commonly found in persons over 50 years old or in certain ethnic groups. To investigate the statistical, rather than physiological basis of such warnings, this study examines correlations globally on a nation-by-nation basis between the statistical data concerning COVID-19 fatalities among the populations of the ninety-nine countries with the greatest number of SARS-CoV-2 infections plus the statistics of potential co-morbidities that may influence the severity of the infections. It examines reasons that may underlie of the degree to which advanced age increases the risk of mortality of an infection and contrasts the risk factors of SARS-Cov-2 infections with those of influenzas and their associated pneumonias.

## 1. Introduction and context

In only a few months after its disclosure by Chinese health authorities, the SARS-CoV-2 virus had spread around the world. By late winter of 2020, the World Health Organization had designated the COVID-19 infection caused by the virus to be a worldwide epidemic. As seen from the effects of the roughly 80 million infections reported by the end of 2020, COVID-19 can manifest in mild, flu-like symptoms or far more seriously as a severe and often deadly respiratory disease with pneumonia.

From the outset of the COVID-19 pandemic, the public has been treated to numerous speculations about the degree to which age and/or various underlying morbidities may amplify the risk of intensifying the severity of infection by the SARS-CoV-2. Authoritative sources such as the U.S. Center for Disease Control (CDC) [Ref.1] have issued warnings. Conditions cited by the CDC as increasing risk include cancer, chronic kidney disease, obesity, coronary disease, Type 2 diabetes mellitus, and sickle cell disease. The CDC also warns that asthma, hypertension, and liver disease among others *might* subject a person to increased risk. In some countries such as the United States, the incidence of COVID-19 has been more prevalent in some ethnic groups than others leading to speculations that that disparity may be biologically rather than behavior based. Such differences are not unknown; for example, sickle cell disease is most commonly found among persons whose ancestors come from Africa and Mediterranean countries where malaria is a prevalent affliction.

As many of the diseases cited by the CDC are more common in persons in late middle age and older, a common warning early in the course of the pandemic was that SARS-CoV-2 presented particular danger to persons over 50 years old. Very early, as in the initial wave of cases in China [2] and in the strong wave of cases in Italy, the probability of death due to COVID-19 was judged to be a strong function of a patient’s age, being only a few percent for those under 50 and rising to nearly 20% for patients over 80. The large number of fatalities [3] in care homes in New York, the United Kingdom and elsewhere have fueled speculations about the risks of co-morbidities frequently seen in the elderly to make contracting COVID-19 fatal. An alternative explanation is the decrease in immune functions with aging. [4]

Why is COVID-19 more dangerous to the elderly than to younger persons? Complicating the answer to this question, the actual mortality rate of COVID-19 remains highly uncertain, as the prevalence of asymptomatic infections has been estimated to be from 2 to 5 times more than infections with clearly defined symptoms. An early exemplary source of testing-based data was provided by the passengers aboard the Princess Line cruise ship, the Diamond Princess on which half of the passengers who tested positive for COVID-19 were asymptomatic or at least pre-symptomatic. [5] To some degree that uncertainty might explain the very wide distributions of reported (or apparent) rate of mortality (case fatality rate) of COVID-19 in countries ranging from < 0.03% (Singapore) to almost 30% (Yemen). Moreover, in most (but not all) countries the integrated average case fatality rate has declined significantly from its high levels seen in March and April of 2020.

For a less anecdotal (and less speculative) assessment of risk factors for serious consequences of COVID-19, a data-driven examination of worldwide national statistics seems to be in order with the goal of identifying strong correlations of mortality due to COVID-19 with other potential co-morbidities and even with ethnically specific biological factors. This manuscript presents a set of calculations of such linear and multivariate correlations.

## 2. Methods of Analysis

The analysis that follows is not based on clinical or physiological considerations but rather on national epidemiological statistics as reported to international authorities. Unless otherwise indicated, the following assumptions underlie the subsequent calculations:

1. *The apparent mortality outcomes (case fatality rates, CFR)* serve as a *reliable proxy* for actual rates of infection, death, and correlation with co-morbidities; we define

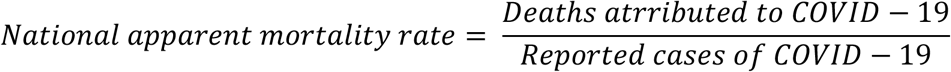 The apparent mortality and case number data used in the following analysis are accurate as of 30 December 2020. This analysis does not and cannot account for any uncertainty due to differing national practices in distinguishing between deaths with COVID-19 and deaths due to COVID-19.
2. The sample of 99 countries across all continents *is representative* of potential correlations between COVID-19 case fatality rates and potential co-morbidities or ethnicity. The number of COVID-19 cases in the countries not included is not statistically significant. Nevertheless, outliers with relatively small statistical significance can skew calculated correlations.
3. *Linear correlations* are examined on the basis of averaged national data for the year 2020. The sources that describe the prevalence of disease are from the World Health Organization^1^, Worldometer^2^, and for economic data the World Bank as reported by Trading Economics.^3^. This analysis *assumes* that the published WHO data concerning the fatalities ascribed to diseases in a given country constitute valid proxies for the prevalence of those maladies in national populations. In the case of obesity, the reported number is the percentage of the population with a BMI exceeding a WHO established standard for a person of that sex.

The study examined the following factors:

*Demographics:* geographical region, population, national median age;

*SARS-CoV-2:* number of COVID-19 tests, confirmed cases of COVID-19 as reported by government authorities, and the apparent case fatality rate;

*Medical factors:* incidence of flu, lung disease, asthma, obesity, heart disease, common cancers, hypertension, chronic kidney disease, diabetes, and malnutrition;

*Economics:* GDP-PPP, average household size, % population living in slums, health expenditures per capita, and WHO universal health coverage index;

*One random (or pseudo-random) variable* in the range from 0 to 100.

Examination of the data begins with computing linear correlations between variables. The evaluation of the linear correlation herein uses the Pearson “product moment correlation” to evaluate linear relationships between data sets:

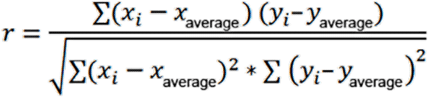

One may estimate the *statistical significance* of calculated correlations by computing *r* for two variables that are *uncorrelated by construction*; i.e., apparent COVID-19 mortality and a random variable in the range from 1 to 100. Once linear correlations have been computed, the next step is evaluating cross-correlations among variables and performing a multivariate analysis.

The 99 countries sampled in this study have been selected as those reporting the largest number of COVID-19 infections. The countries listed in Table 1 represent five geographical regions: the Americas, Asia, Europe, Africa, and Middle East plus Central Asia. Regional populations are included. The combined population of nearly 5.5 billion persons accounts for the strong preponderance of all cases reported worldwide. The data cutoff date was December 30, 2020.

**Table 1.** Countries sampled grouped into five regions. As Yemen is a statistical outlier in apparent mortality, many plots omit its data point for visual clarity.

| Americas<br>*pop = 977 M* | Asia<br>*pop = 2504 M* | Europe<br>*pop = 725 M* | Africa<br>*pop = 768 M* | Middle East<br>*pop = 487 M* |
| --- | --- | --- | --- | --- |
| *Argentina* | *Australia* | *Albania* | *Algeria* | *Afghanistan* |
| *Bolivia* | *Bangladesh* | *Armenia* | *Cameroon* | *Bahrain* |
| *Brazil* | *China* | *Austria* | *Congo* | *Egypt* |
| *Canada* | *India* | *Azerbaijan* | *Ethiopia* | *Iran* |
| *Chile* | *Indonesia* | *Belarus* | *Ghana* | *Iraq* |
| *Columbia* | *Japan* | *Belgium* | *Ivory Coast* | *Israel* |
| *Costa Rica* | *Kazakhstan* | *Bosnia* | *Kenya* | *Lebanon* |
| *Dominican Republic* | *Kyrgyzstan* | *Bulgaria* | *Libya* | *Kuwait* |
| *Ecuador* | *Korea* | *Croatia* | *Madagascar* | *Oman* |
| *El Salvador* | *Malaysia* | *Czechia* | *Mali* | *Qatar* |
| *Guatemala* | *Nepal* | *Denmark* | *Morocco* | *Saudi Arabia* |
| *Honduras* | *New Zealand* | *Estonia* | *Nigeria* | *Turkey* |
| *Mexico* | *Pakistan* | *Finland* | *South Africa* | *UAE* |
| *Panama* | *Philippines* | *France* | *Sudan* | *Uzbekistan* |
| *Paraguay* | *Singapore* | *Germany* | *Uganda* | *Yemen* |
| *Peru* | *Thailand* | *Greece* | *Zambia* |  |
| *USA* | *Taiwan* | *Hungary* |  |  |
| *Venezuela* |  | *Ireland* |  |  |
|  |  | *Italy* |  |  |
|  |  | *Macedonia* |  |  |
|  |  | *Moldova* |  |  |
|  |  | *Netherlands* |  |  |
|  |  | *Norway* |  |  |
|  |  | *Poland* |  |  |
|  |  | *Portugal* |  |  |
|  |  | *Romania* |  |  |
|  |  | *Russia* |  |  |
|  |  | *Serbia* |  |  |
|  |  | *Spain* |  |  |
|  |  | *Sweden* |  |  |
|  |  | *Switzerland* |  |  |
|  |  | *Ukraine* |  |  |
|  |  | *UK* |  |  |

The SARS-CoV-2 related data are sex-aggregated because many countries still do not report sex-disaggregated data (or made these data available publicly). Therefore, the frequently reported sex-based disparities in contagion and in CFR could not be examined with respect to sex-based differences in occurrences of potential co-morbidities.

Figure 1 plots the CFR and the random number that are uncorrelated by construction. The Pearson coefficient for this set of 99 values is −5.3%.

**Figure 1.**
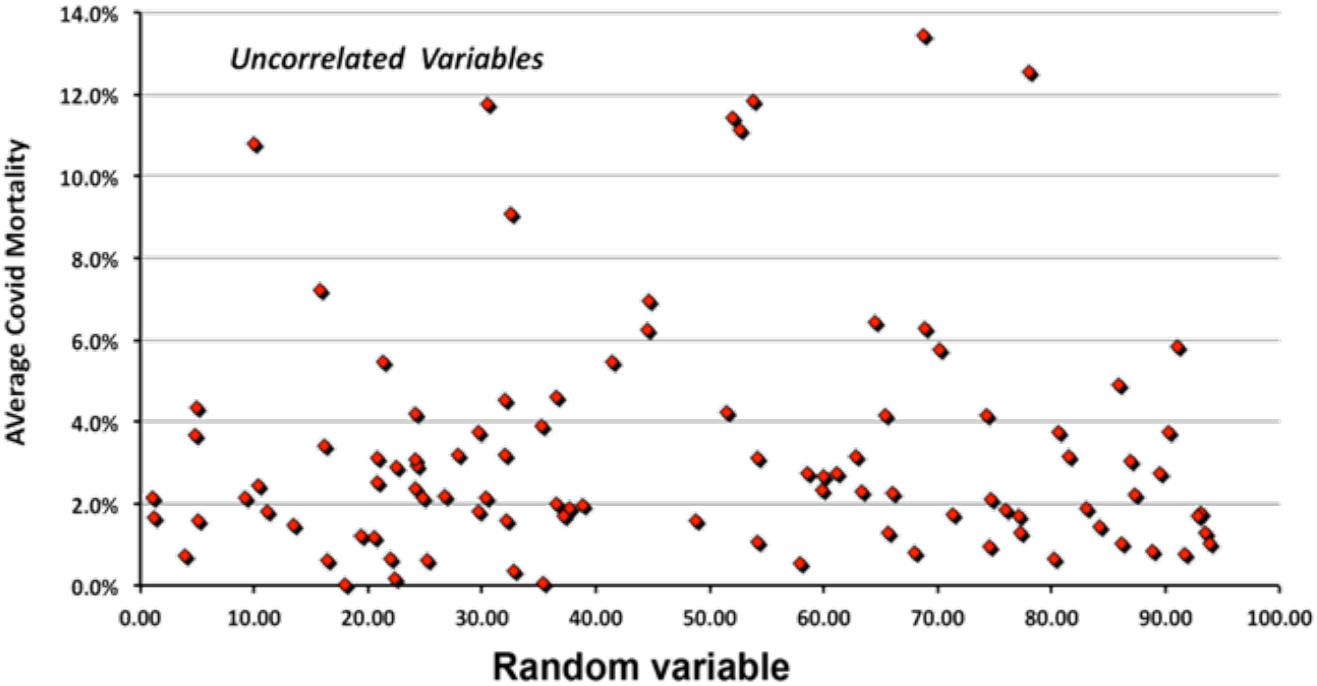
A plot of the variables uncorrelated by construction.

A potential limitation of this approach is that all mortality data have equal weight in the calculation of correlation. One check of whether this Ansatz introduces a bias is the correlation between apparent national mortality rates and national populations. Doing so, yields a −1.4%, a value close to the Pearson coefficient for uncorrelated variables. Another possible way to attribute a rational weighting is plotting the variation of COVID-19 deaths per capita against the possible risk factor. However, the number fatalities per capita depends strongly on national public health policies, on national efforts to prevent spread of the SARS-CoV-2 virus, on GDP, and other non-medical considerations. Differences in COVID-19 statistics between Norway and Sweden are cases in point.

## 3. Examination of linear correlations

To gain confidence in this statistical approach one can plot two variables for which one may expect to see a correlation (Figure 2). Here the linear correlation is quite high, 62.5%. Examining Fig. 2 more closely suggests a limitation of the considering only linear correlations. The countries circled in red show a strong correlation while those in the green circle that show scarcely any correlation of a nation’s wealth with the age of its population. Clearly, a refinement of the statistical approach is needed. By identifying the data underlying each point with each country’s region reveals that median age and national wealth are essentially uncorrelated for European nations but strongly correlation for countries in Africa and Asia. *Regional grouping* was thus adopted throughout this study.

**Figure 2.**
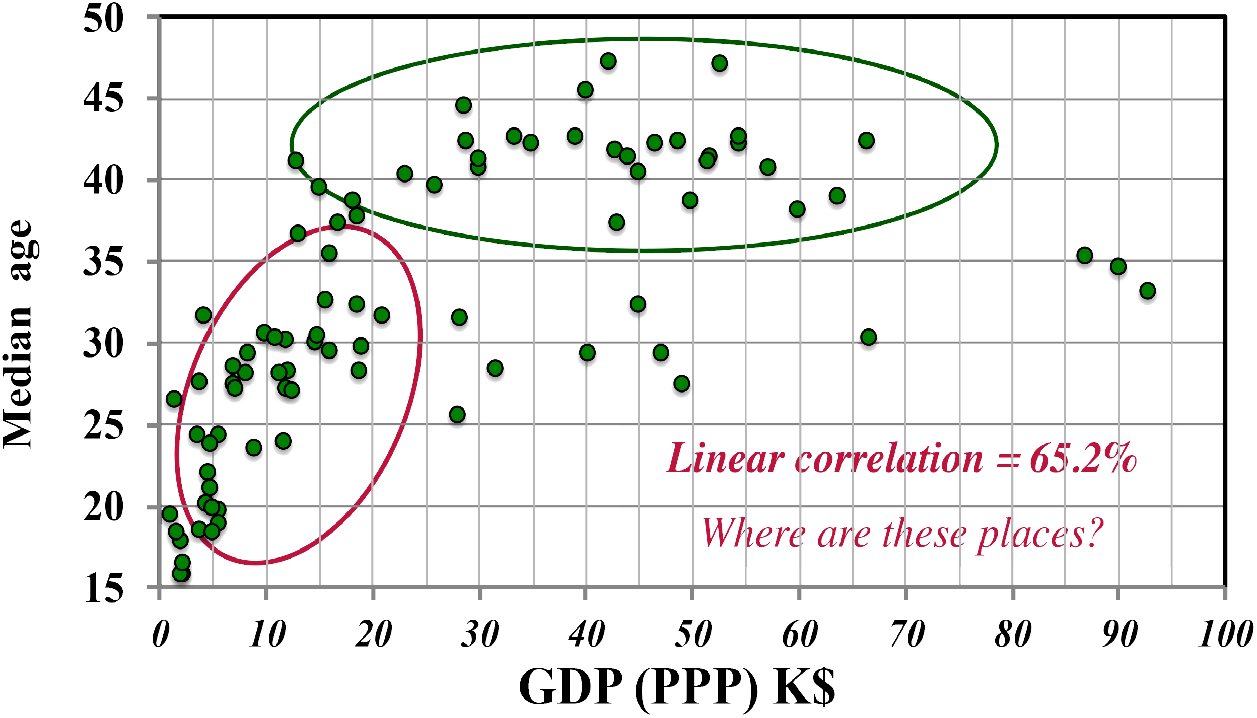
The plot of GDP-PPP corrected for purchasing power^4^ versus median age.

To illustrate the utility of this refinement, Figure 4 shows the relation between deaths per 100k persons due to malnutrition as a function of national wealth measured by GDP corrected for purchasing power. The relatively strong (45.5%) correlation is driven by the high rates of malnutrition in Africa, Central America, and the poorer countries of Asia. No such effect is apparent in Europe.

**Figure 3.**
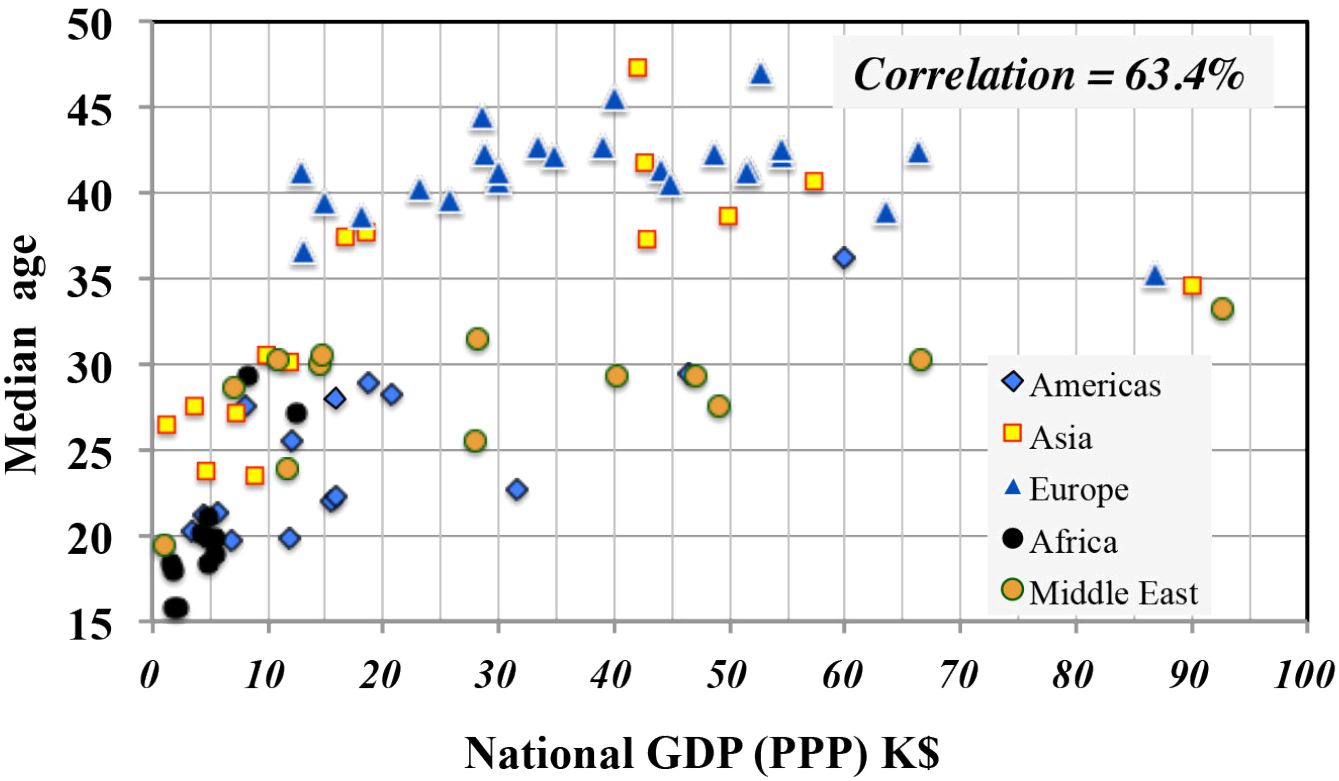
GDP vs. Median age in countries from the five regions under study

**Figure 4.**
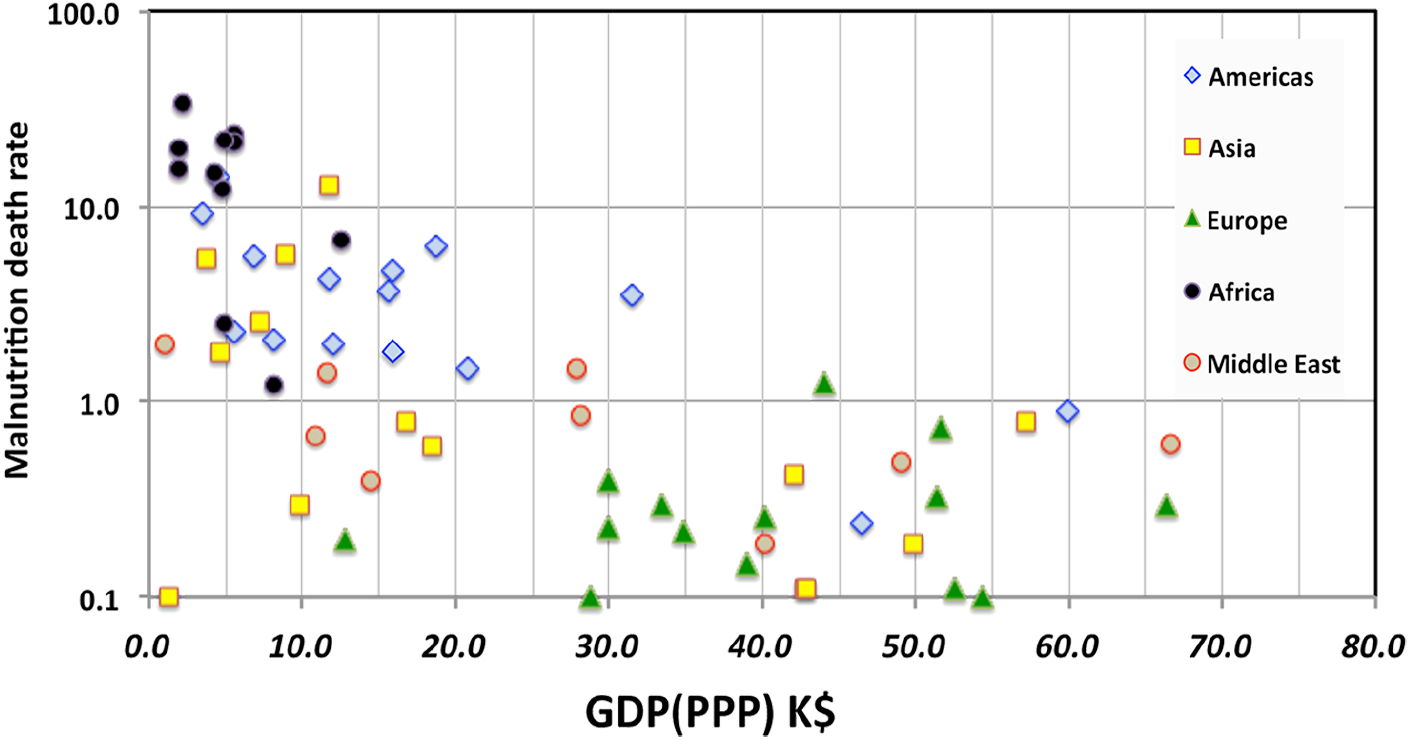
It is not surprising that poverty is correlated with malnutrition.

From the outset of the pandemic, national health authorities have warned the public about the increased risk of mortality for persons 60 years old and older. Figure 5 shows an example of the basis for such warnings in the data provided by the UK Office of National Statistics in September 2020 [5]. Again one asks why should the severity of a COVID-19 infection be a function of age?

**Figure 5.**
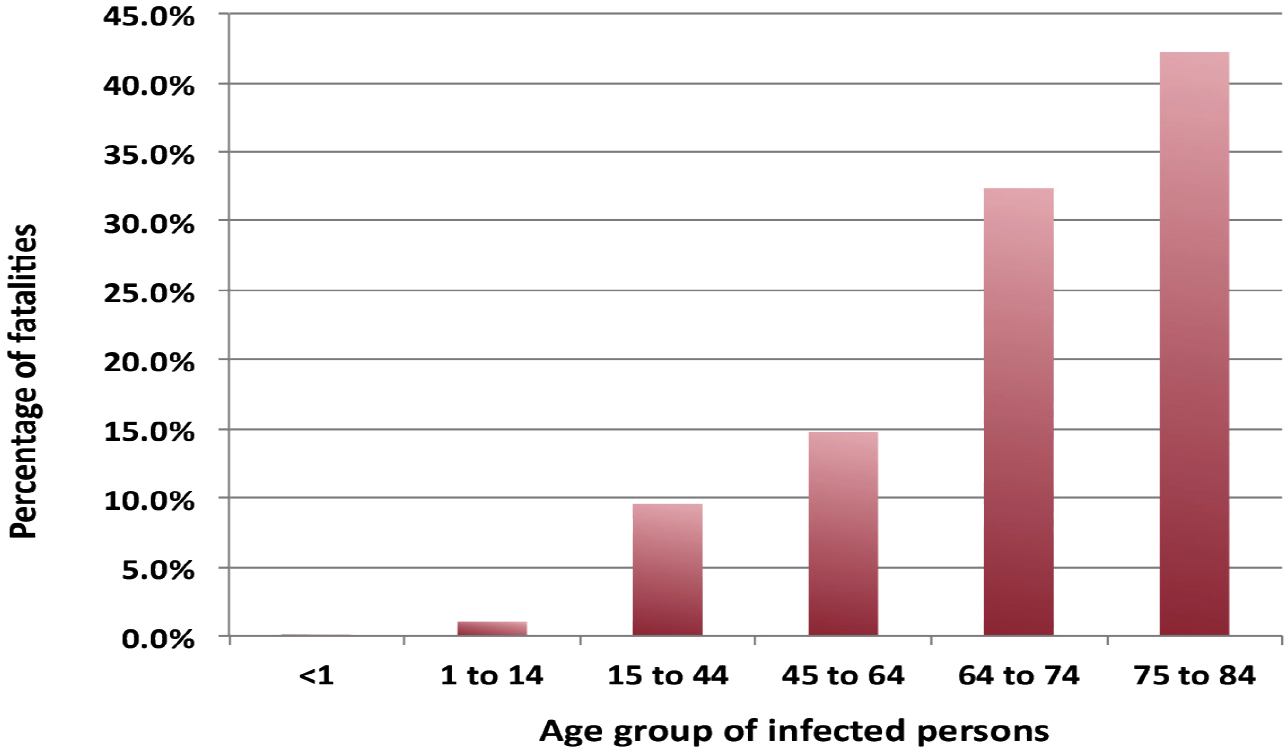
Deaths attributed to COVID-19 by the UK Office of National Statistics.

From such data, one might expect a very strong correlation between the national apparent CFR and the median age of a country’s population. Even accepting the hypothesis of universality for the data of Figure 5, one should first multiply these rates by the demographics of a nation’s population normalized to the U.K. population grouped into the same age bins. Such a plot (in Figure 6a) shows a surprising result. The overall linear correlation is negative, −18.1%, partially due to the disparity among the regions: −25.8% for the Americas, 5.2% for Asia, 14.1% for Europe, 2% for Africa and −60.8% for the middle East and Central Asia.

**Figure 6(a).**
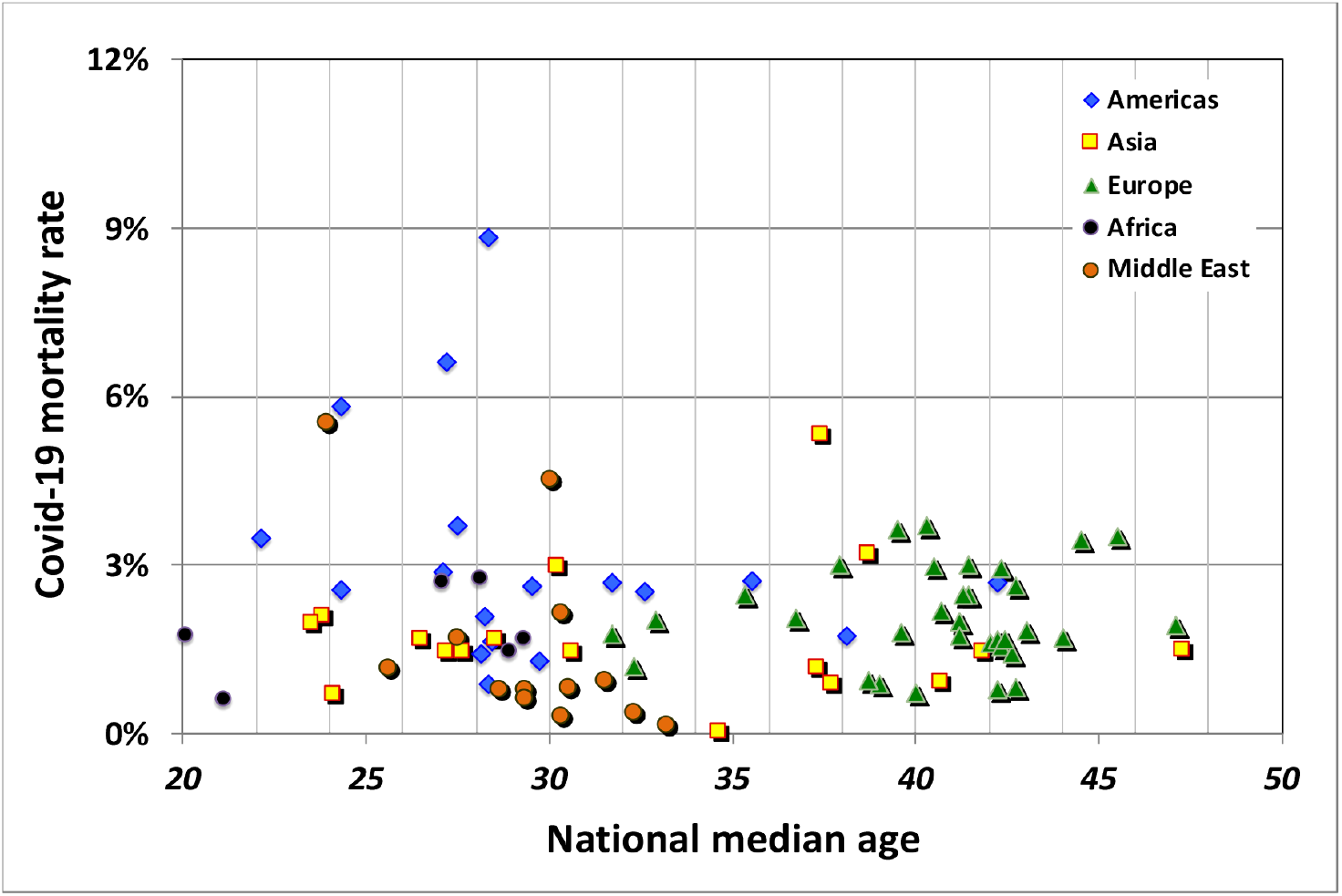
National median age versus CFR for the five regions

Instead of plotting the COVID-19 CFR versus national median age, one can examine the dependence on the percentage of the population of age 65 or greater (figure 6b). The overall correlation (−8.1%) is negative, consistent reference [5], but is mostly the result of regional variations with a larger, but still relatively low correlation (∼19%) in Europe and Africa.

**Figure 6.(b).**
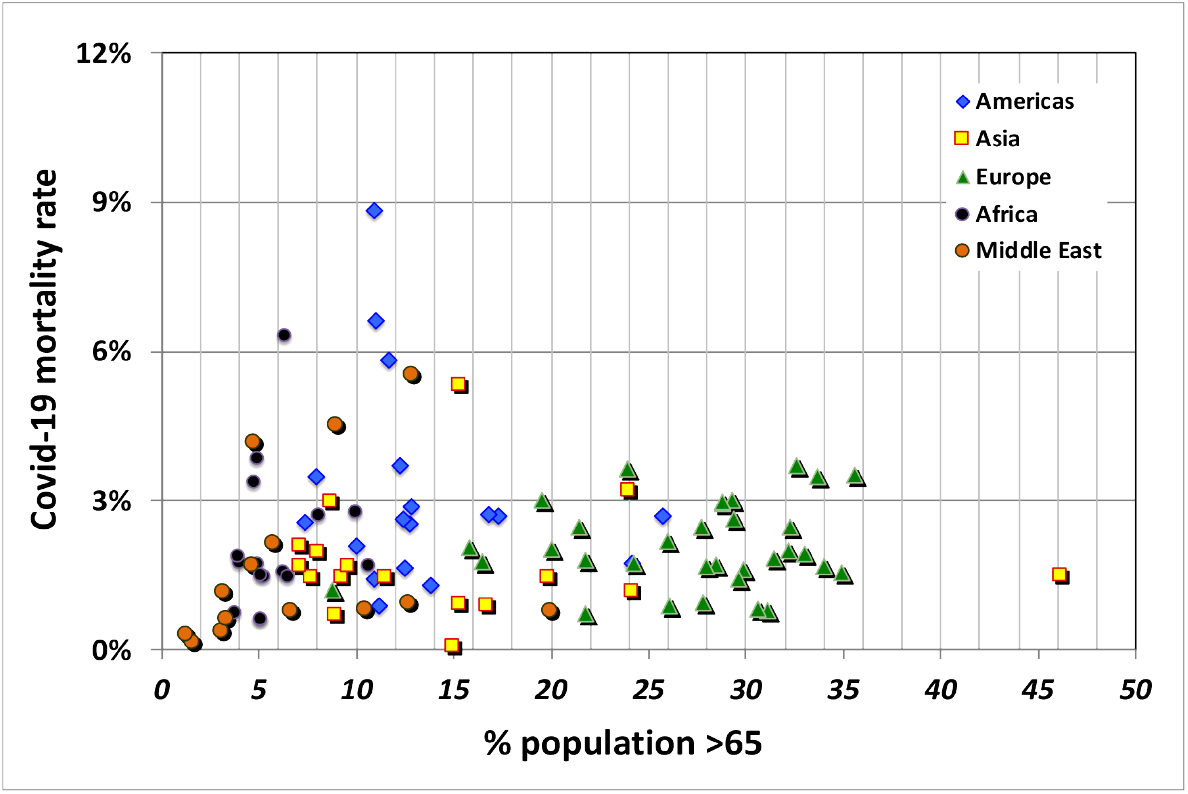
Percentage of population over 65 versus apparent mortality.

As a measure of the influence of the age of a population on SARS-CoV-2 contagion, the national rate of confirmed cases of COVID-19 per 1M persons with respect to the percentage of population older than 65 (Figure 7) displays a moderate correlation of 44.7%.

**Figure 7.**
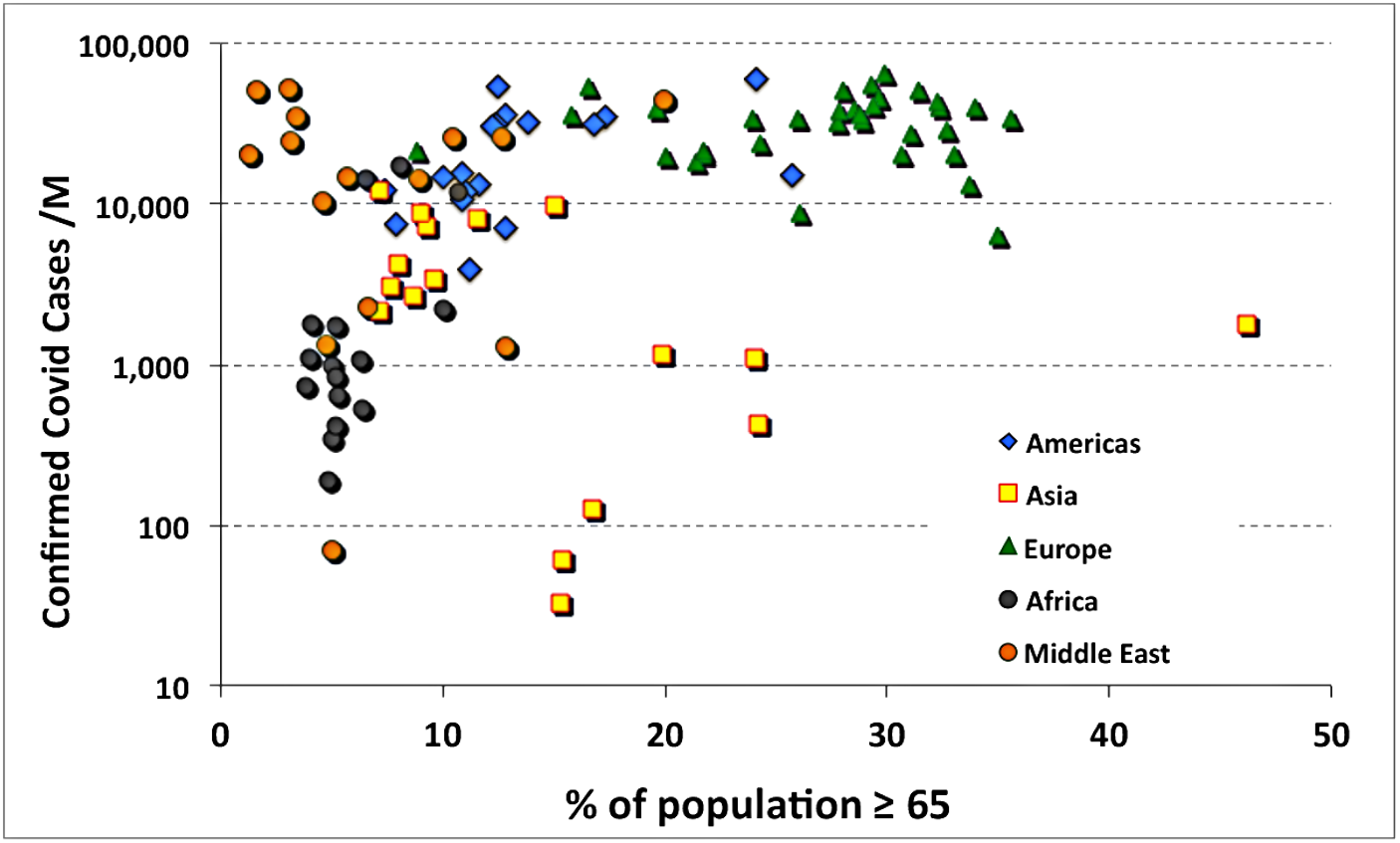
Confirmed COVID-19 cases as a function percent of population older than 65.

One may hypothesize that the “care home effect,” i.e., the large numbers of deaths seen in nursing homes in Italy, the U.K. and N.Y. was more the result of poor hygienic practices plus the generally weakened physical state and reduced immune function of residents than by any extreme dependence of the lethality of COVID-19 infections on specific underlying disorders. That hypothesis is investigated in the following analysis. The linear correlations of age with various potential causal factors shown in Figure 8 suggest candidates to examine to explain the “care home effect.”

**Figure 8.**
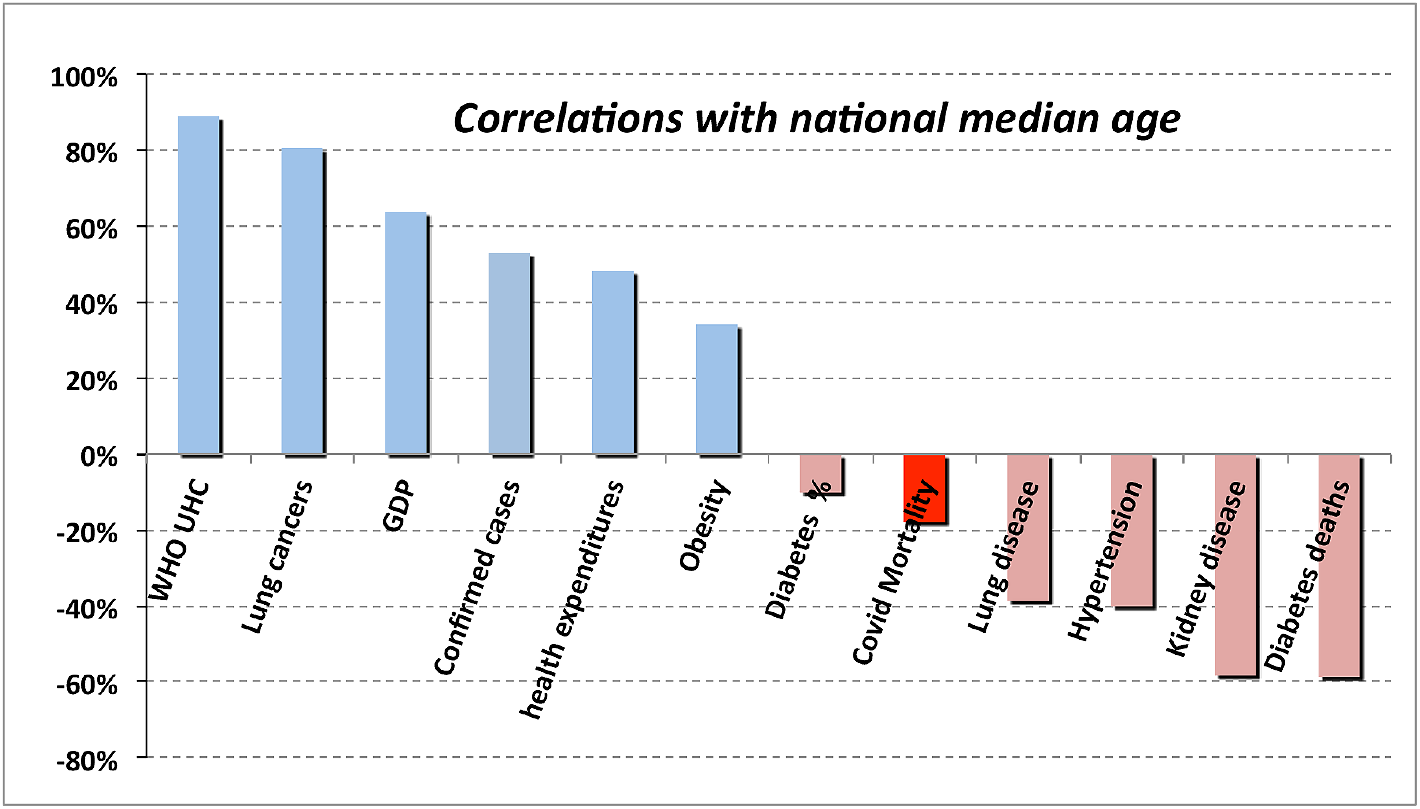
Correlations of potential risk factors with median age

In addition to particular underlying factors, the “care home effect” also reflects a generally very weakened physical condition of many occupants of care homes that would render any pneumonia-inducing disease lethal. Finally, the data shown in Figure 8 show no evidence that age alone influences the probability of a person becoming infected by the SARS-Cov-2 virus.

The data of Figure 8 also explain what may seem like a starting result, namely that the correlation of COVID-19 CFR with the age of national populations is negative globally. That is explained by the very strong correlations between national median age and adjusted GDP(64%), health care expenditures (48%) and the WHO Universal Health Care Index. The nations with the oldest populations are generally those that are the wealthiest and in which health care services are the largest, thus reducing the level of mortality.

As suggested by the results in Figure 8, an example (Figure 9) illustrates the utility of the statistical approach used herein. In contrast with infections due to SARS-Cov-2, the incidence of death from influenza-induced pneumonia is highly correlated (−65.2%) with the median age of the population. The correlation also displays a strong regional dependence. The correlation is negative for the same reasons as previously explained for COVID-19.

**Figure 9.**
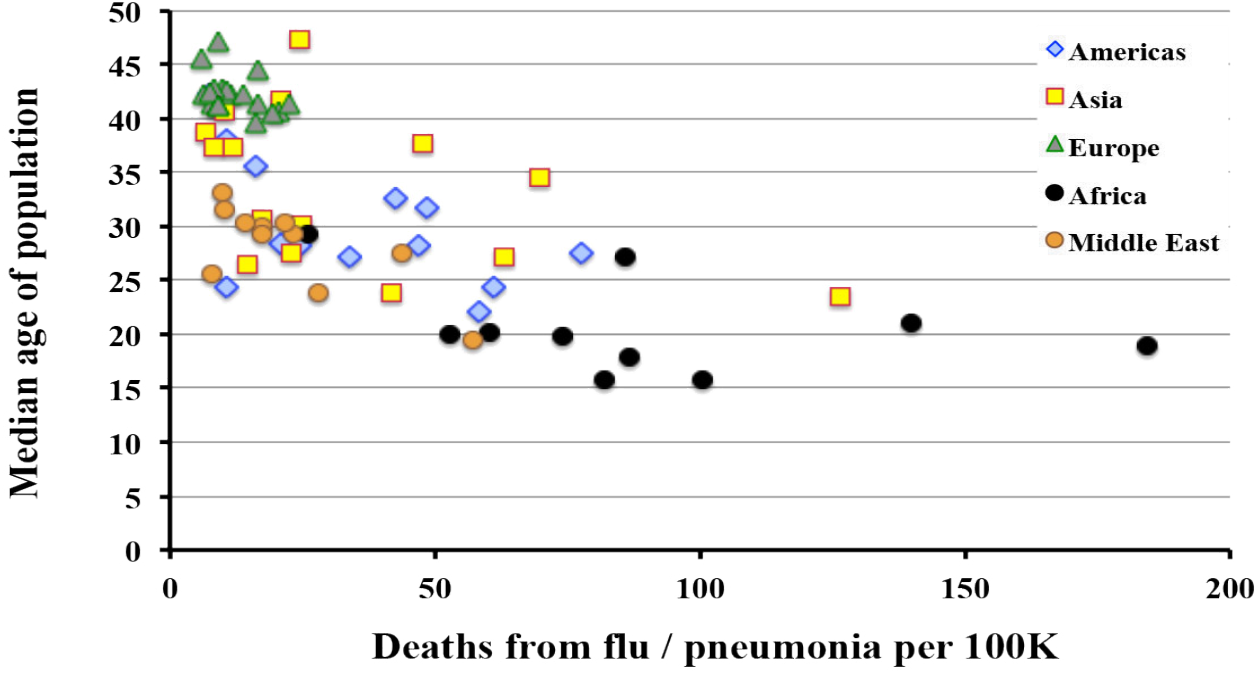
Incidence of flu related pneumonia deaths as a function of national median age.

As the COVID-19 often presents as a severe respiratory disease and strengthened by the results for influenza in Figure 9, one asks whether the severity of COVID-19 infections is correlated with incidence of asthma. As displayed in Figure 10, the global value is small but not negligable, 16.5%, largely driven by the correlation (68.1%) in the Middle East.

**Figure 10.**
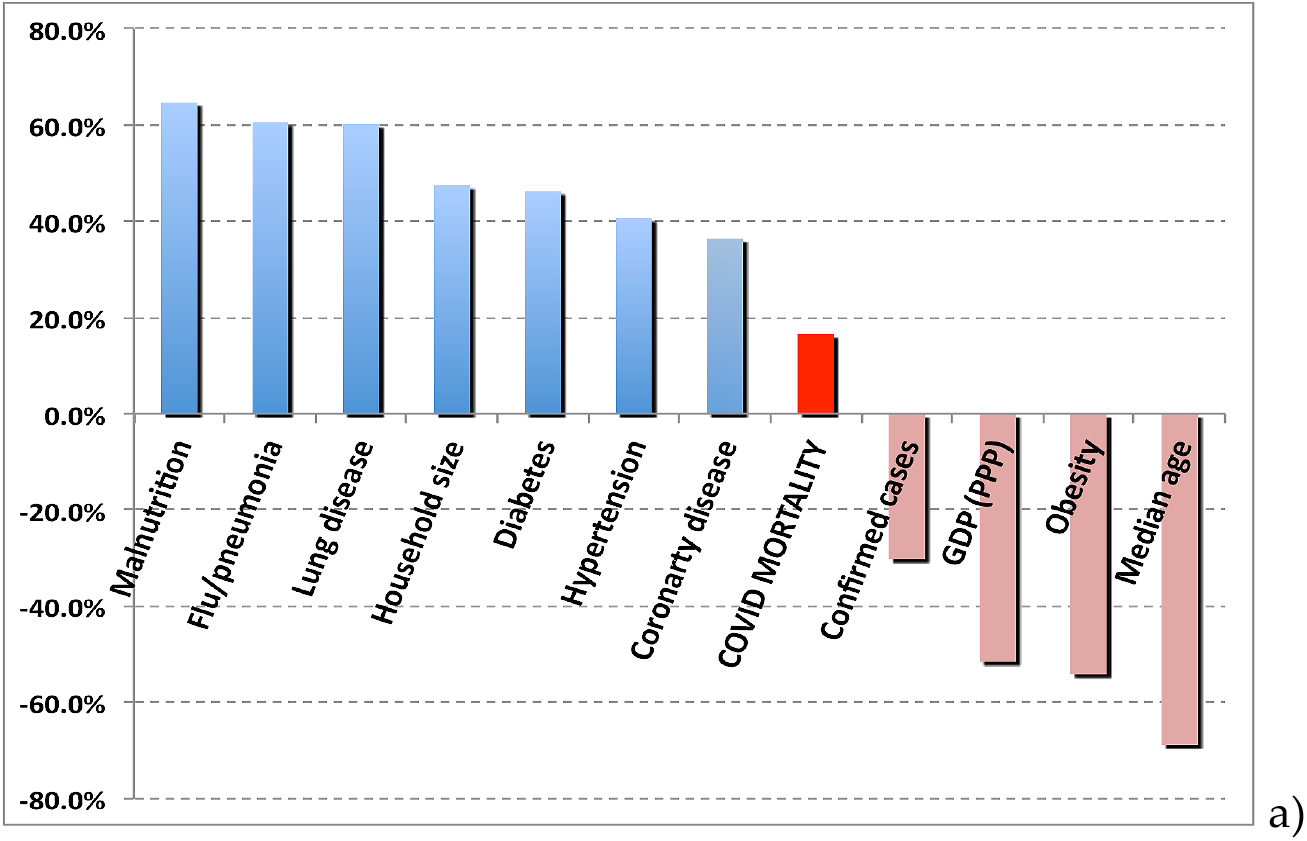
a) Correlation of severe asthma with COVID-19 CFR

For asthma as a co-factor, the contrast with influenza related pneumonia (Figure 11) is striking. The relatively high, overall correlation of 59.4% is seen in all regions. Any reference to COVID-19 as a “flu-like” infection or as a “superflu” is grossly misleading.

**Figure 11.**
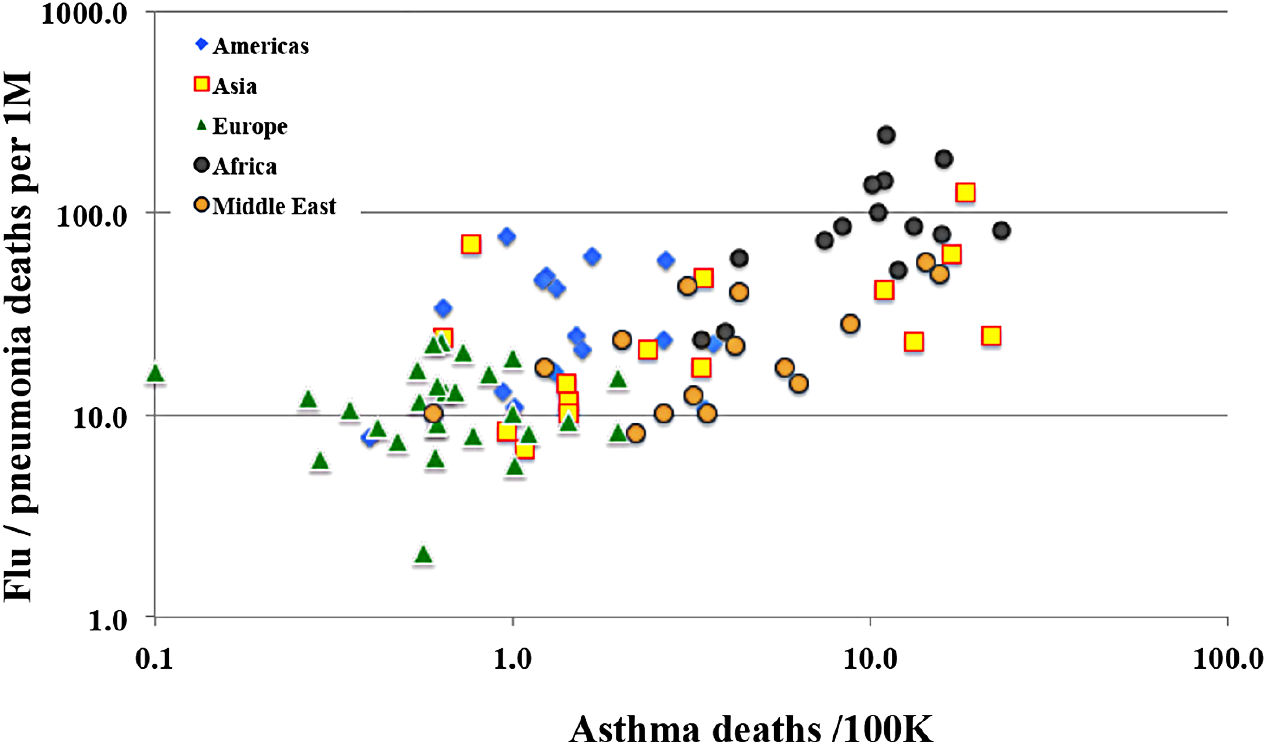
The incidence of death from flu related pneumonia is relatively strong especially in developing countries.

An early warning of the U.S. Centers for Disease Control was that obesity represented an underlying co-factor that could lead potentially to severe consequences of an COVID-19 infection. However, once again actual the national data of (Figure 12) display essentially no correlation (−1.7%) of COVID-19 CFR with the percentage of a country’s population considered obese.

**Figure 12.**
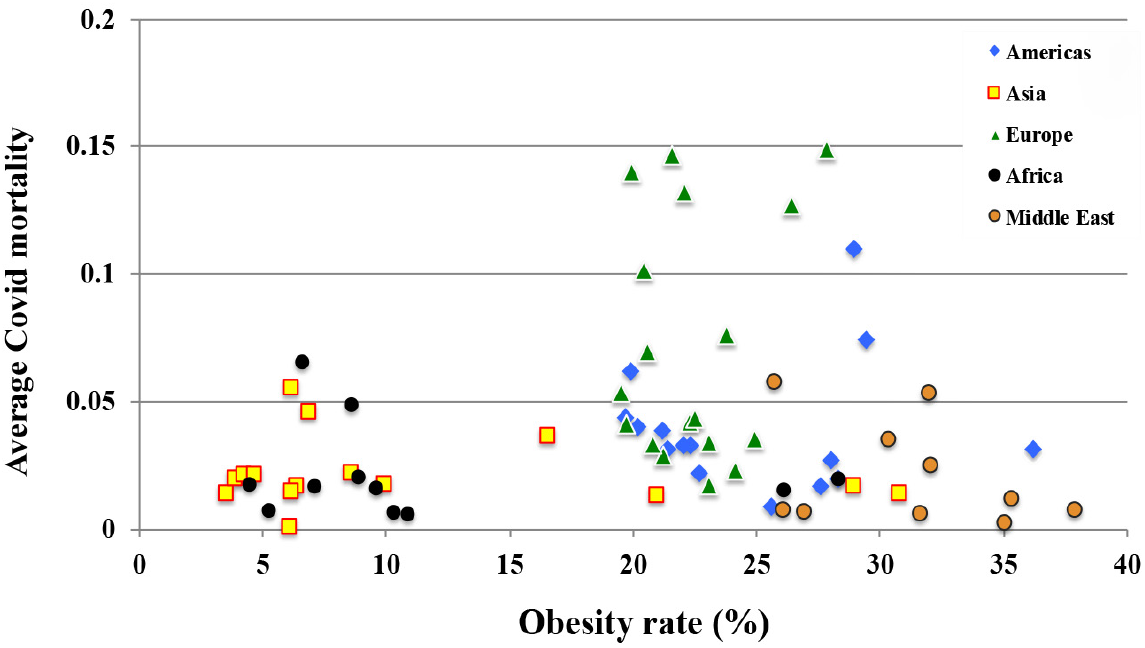
The correlation of national incidence of obesity with apparent national rates of COVID-19 mortality. The minimal correlation is characteristic of all regions.

On might suggest that a better metric of obesity in a country is the average Body Mass Index (BMI in kg/m^2^) of its population. Substituting BMI for the metric of Figure 11, the correlation increases to 5.2%, still very small. Moreover, that figure may be misleading comparing region to region as the correspondence between BMI and Body Fat percentage varies considerably (10% to 20%) from country to country.

The contribution of obesity to the *outcome* of other pulmonary disorders is significantly different to that of COVID-19 as is displayed in Figure 13. Curiously, obesity has a significant correlation (51.6%) with the risk of contracting infection from SARS-Cov-2, although not with the apparent outcome of the infection. The observation of increased risk of infection (although not its outcome) has been previously reported in [6]. Reference [7] reports an increased risk of infection (32.9%) for people with chronic kidney disease. That correlation is not seen in the statistics of this study which has consistently found an temporally increasing negative correlation.

**Figure 13.**
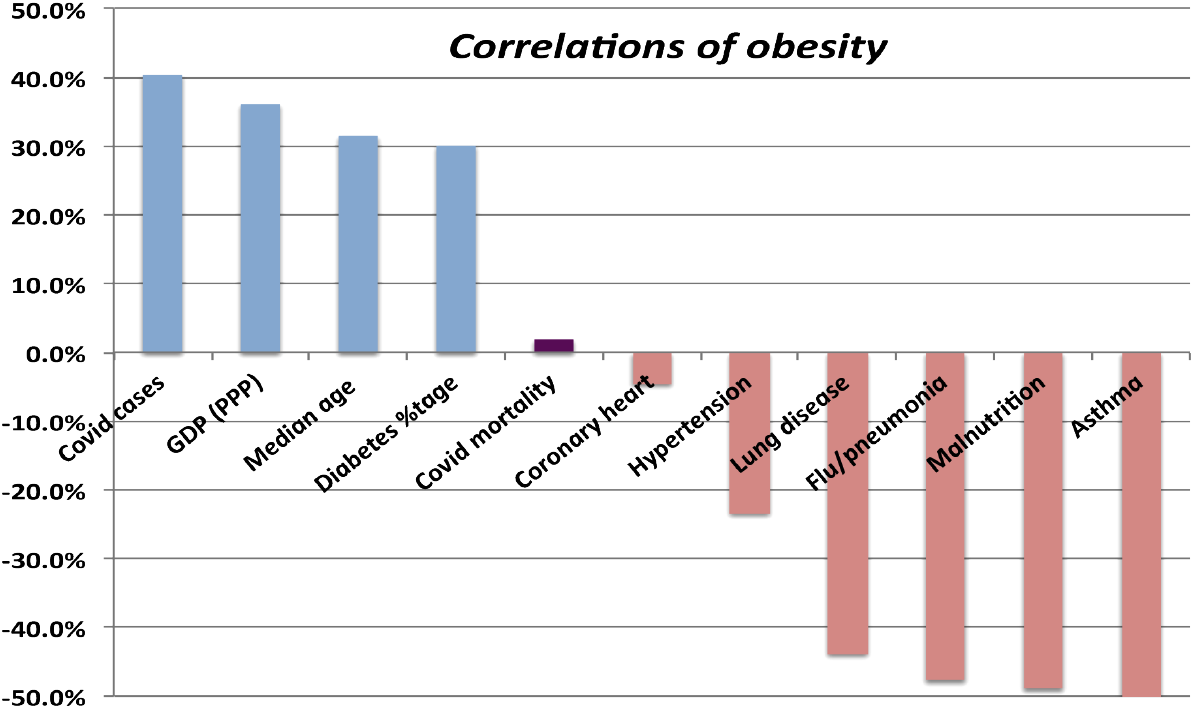
Correlations of obesity rates with COVID-19 mortality and other conditions. For most of conditions, rates are based on deaths per 100K persons.

One might speculate that as a chronic respiratory disorder involving pulmonary airways, asthma may increase the seriousness of consequences of COVID-19 and its induced pneumonias; however, Figure 14 shows no such significant correlation (5.3%). Examining the correlation of COVID-19 mortality with other lung diseases (Figure 15) also shows a tiny correlation (1.3%). In contrast, the relationship of influenza-induced pneumonias with asthma and other lung diseases presents a correlation that is quite high, 59.4% and 34.8% respectively.

**Figure 14.**
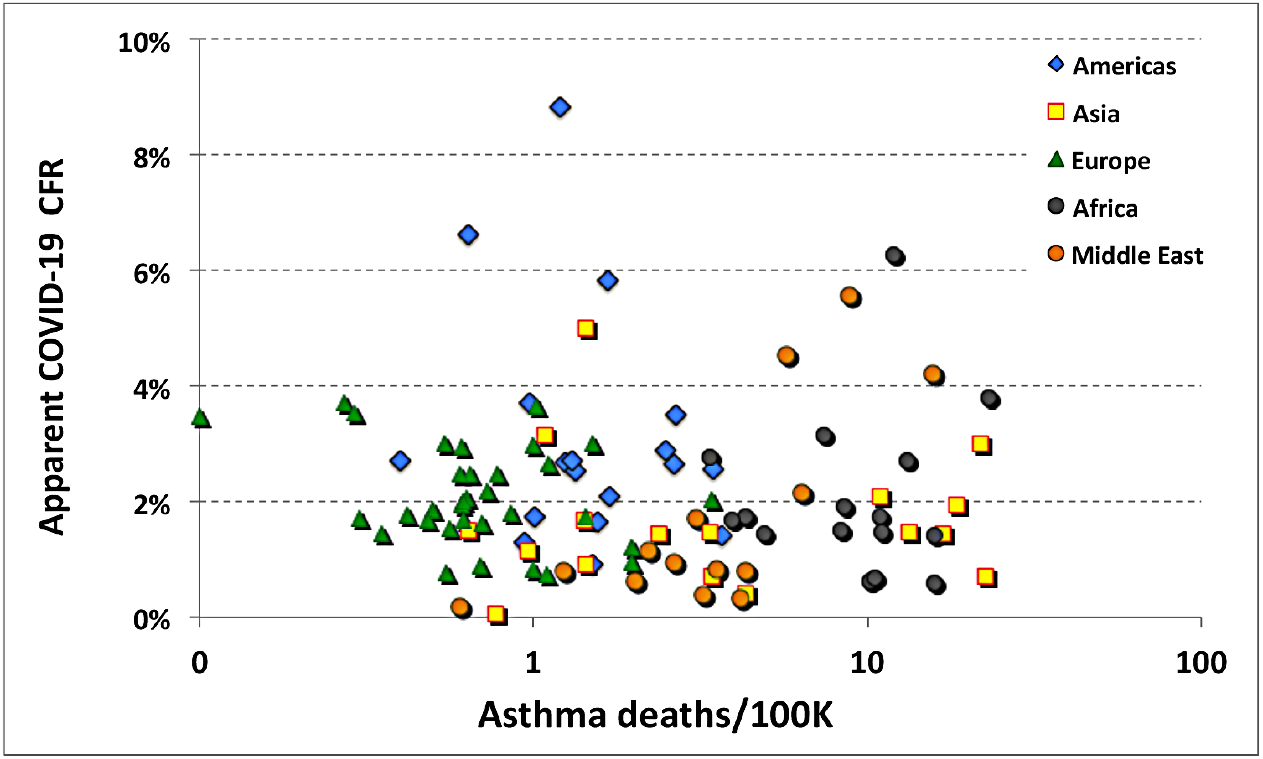
Correlation of asthma with apparent COVID-19 mortality rate.

**Figure 15.**
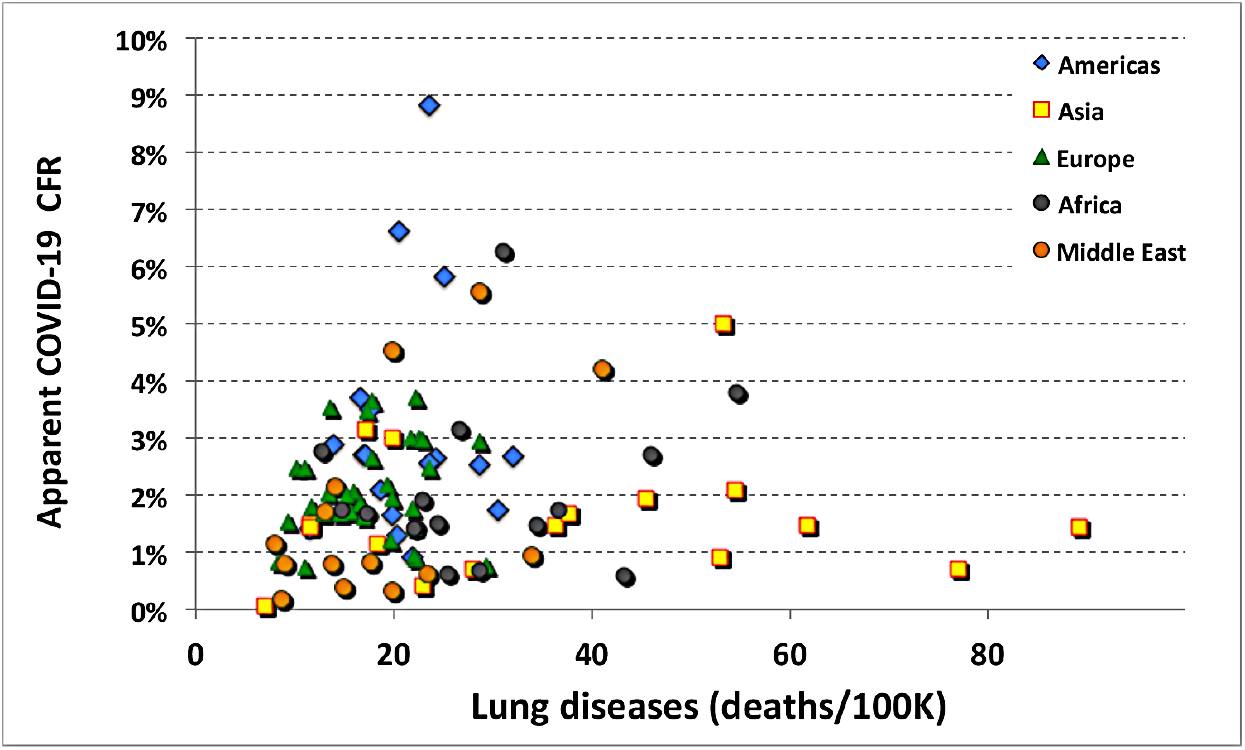
Correlation of COVID-19 mortality with the incidence of lung disease.

With respect to their effects on patients with underlying conditions, influenza and COVID-19 are very different diseases.

Another early warning to persons with underlying conditions concerned diabetes mellitus. That suspicion is echoed by the strong dependence with age shown in Figure 8. Whether one measures the incidence of diabetes by deaths due to diabetes or to the reported national rates of diabetes in adults (20 to 79 years of age), the correlation with COVID-19 mortality is similarly low (10.9%). In otherwise healthy persons, diabetes does not appear to be a significant risk factor with respect to the serious complications of infection by SARS-CoV-2.

Figure 16 and Table 2 summarize the linear correlations of COVID-19 CFR with underlying medical and economic conditions (in green) considered herein.

**Table 2.** Correlation with national values of apparent COVID-19 CFR

| Potential co-factor | Date of COVID-19 CFR statistics |  |  |
| --- | --- | --- | --- |
|  | 30-Dec | 20-Nov | 16-Oct |
| Kidney disease | 26.9% | 28.9% | 17.6% |
| household size | 22.5% | 22.8% | 12.6% |
| heart disease | 20.4% | 19.4% | 9.9% |
| Asthma | 16.5% | 16.8% | 9.1% |
| diabetes deaths | 13.3% | 14.8% | 5.0% |
| covid deaths per 1M | 9.2% | 7.0% | 17.0% |
| %slums | 9.0% | 7.2% | 5.9% |
| hypertension | 5.1% | 4.9% | -1.1% |
| flu/pneumonia | 3.4% | 4.0% | -2.0% |
| malnutrition | 1.7% | 0.2% | -3.7% |
| lung disease | 1.3% | 2.4% | -11.2% |
| Random number | 0.9% | -2.4% | 2.6% |
| Diabetes (% population) | 0.9% | 4.6% | -4.3% |
| population | -1.3% | 0.0% | -1.4% |
| obesity % population | -1.7% | -0.6% | 1.4% |
| % population ≥65 | -8.1% | -10.3% | 2.8% |
| covid cases /1M | -15.3% | -16.3% | -8.6% |
| health care expenditure | -15.5% | -14.3% | -2.0% |
| lung cancers | -15.9% | -17.9% | -9.8% |
| life expectancy | -16.3% | -15.2% | -5.5% |
| median age | -17.9% | -19.1% | -7.4% |
| WHO UHC index | -19.7% | -16.8% | -7.6% |
| % population in cities | -19.7% | -17.7% | -12.1% |
| adjusted GDP | -23.0% | -21.5% | -11.9% |
| covid tests per 1M | -28.7% | -25.7% | -11.1% |

**Figure 16.**
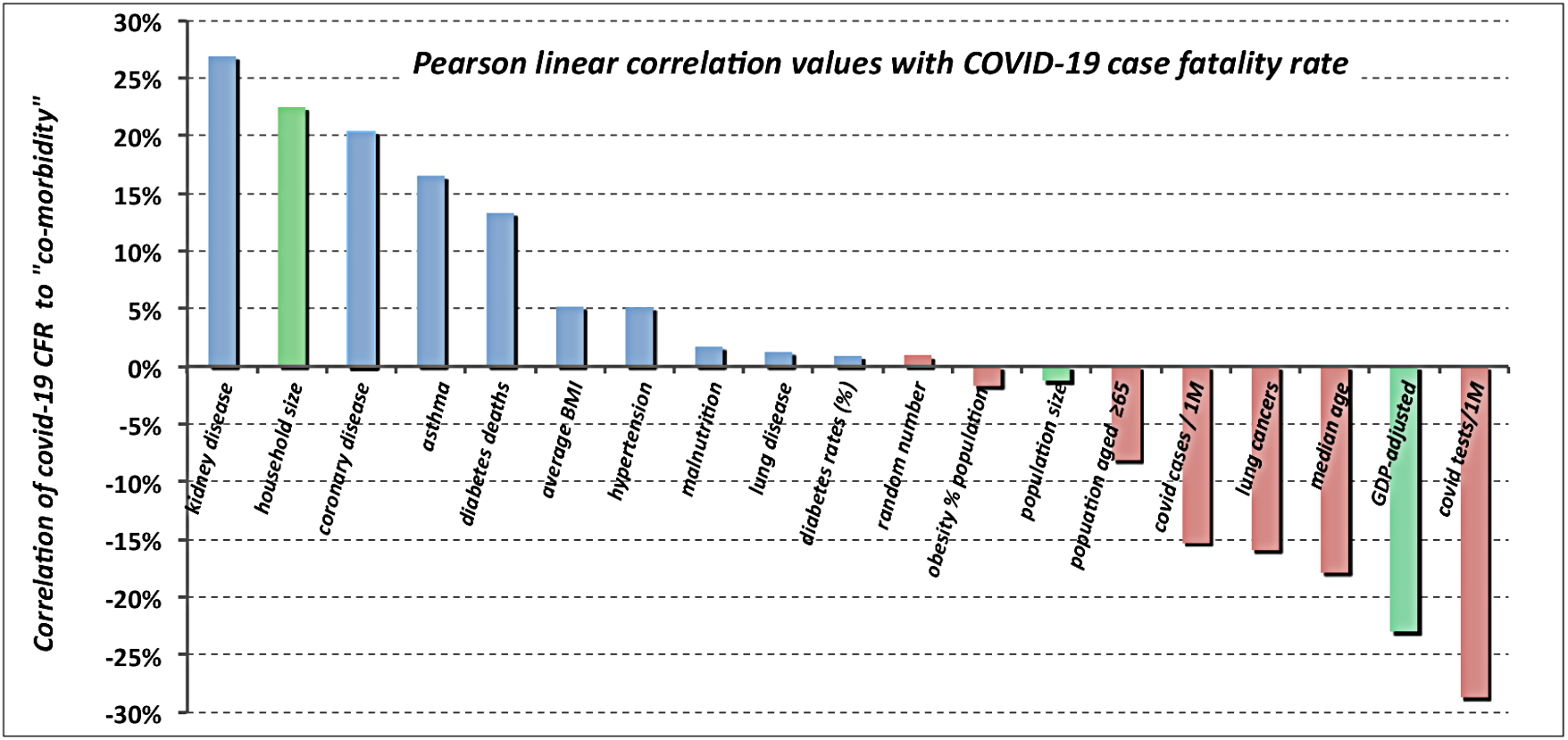
Summary of linear correlations with national COVID-19 CFR data.

As the shown in Table 2, the percentage of population over 65 years of age correlated at best weakly with apparent COVID-19 case fatality rate. One may surmise that poor health care management played a very large role in the “care-home effect.”

One notes in figure 16 the strong negative correlation of CFR with both COVID tests per million and with the number of cases per million. More tests mean earlier detection, more detection of mild and weakly symptomatic cases, better triage and earlier and more effective clinical treatments.

## 4. Cross-correlations and multivariate analysis

Before investigating cross-correlations to search for root causes, one should perform a multivariate analysis of COVID-19 CFR against a common trio of risk factors commonly found in patients in nursing and convalescent homes– namely diabetes mellitus, hypertension, and coronary disease (DHC). For that trio, the coefficient of multiple correlation is 17.1%, not negligible but unlikely to be the root cause of the “care home effect.” Computing the correlation of DHC with deaths due to influenza and its associated pneumonia yields a stronger correlation of 35.9%. Replacing hypertension with asthma in the DHC trio reduces the coefficient of multivariate correlation for COVID-19 mortality to 12.1%. In contrast, analogous analysis for influenza increases the multiple correlation coefficient to 62.7%, demonstrating once again (see Table 3) that influenza and COVID-19 are very different diseases.

**Table 3.** Multivariate correlations for a trio of input variables: namely, diabetes mellitus, hypertension, and coronary disease.

| Output Variable | Regression coefficient | Pearson r values |
| --- | --- | --- |
| COVID-19 | 12.3% | 3.5%, 5.3%, -4.1% |
| Influenza / pneumonia | 43.9% | 38.6%, 14.7%, 24.7% |

Other calculations of multivariate correlations with the apparent national mortality rates of COVID-19 are presented in Table 4.

**Table 4.** Multivariate correlations with national COVID-19 mortality data

| Multiple Variables | Regression coefficient | Pearson r values |
| --- | --- | --- |
| GDP and household size | 7.0% | -5.9%, 5.6 % |
| Obesity and diabetes | 3.5%% | 3.5%, - 7.1% |
| Influenza and lung disease | 11.7% | -6.4%, -14.8% |
| Diabetes, heart and hypertension | 12.3% | 3.5%, 5.3%, -4.1% |
| Median age and # of cases | 13.8% | 0.4%, -13.7% |
| Flu deaths and diabetes | 10.7% | -6.4%, 3.5% |
| Flu deaths and hypertension | 6.8% | -6.4%, -4.1% |
| Obesity, asthma and diabetes | 14.2 % | 3.5%, 5.3%, 3.5% |

### 4.1. Cross-correlations

The previous section argues and Figure 17 illustrates the striking contrast between the correlations of COVID-19 with those of influenza/pneumonia with respect to other potential underlying conditions.

**Figure 17.**
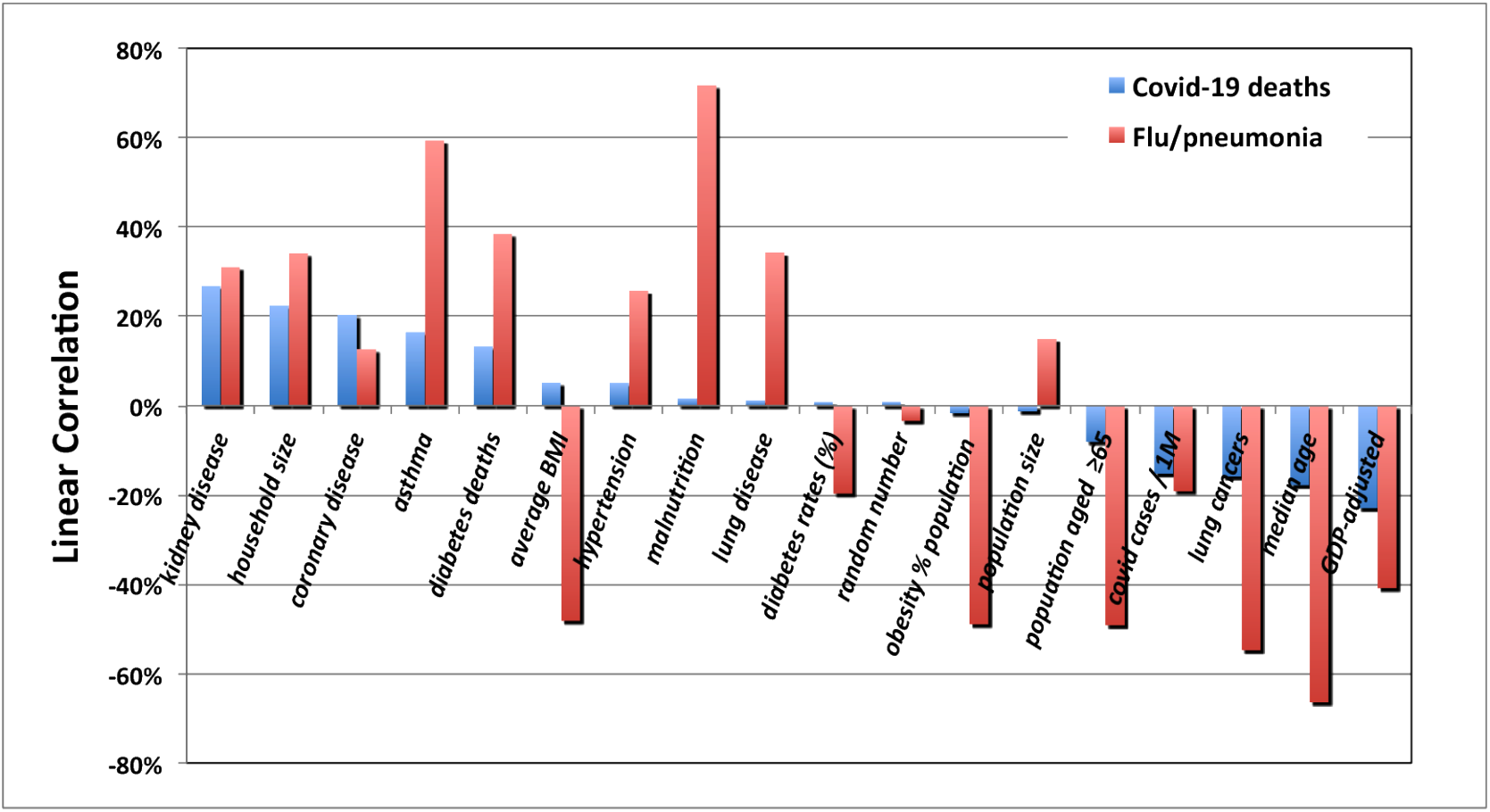
Contrast between correlations of COVID-19 with of flu-induced pneumonia

An examination of cross-correlations (Figure 18) is best displayed in plots ordered in strength of influence of given conditions considered for their possible correlation with the outcome of SARS-CoV-2 infections. In order of presentation, these factors are national median age (Figure 8), obesity (Figure 13), asthma (Figure 18), and diabetes mellitus (Figure 22).

**Figure 18.**
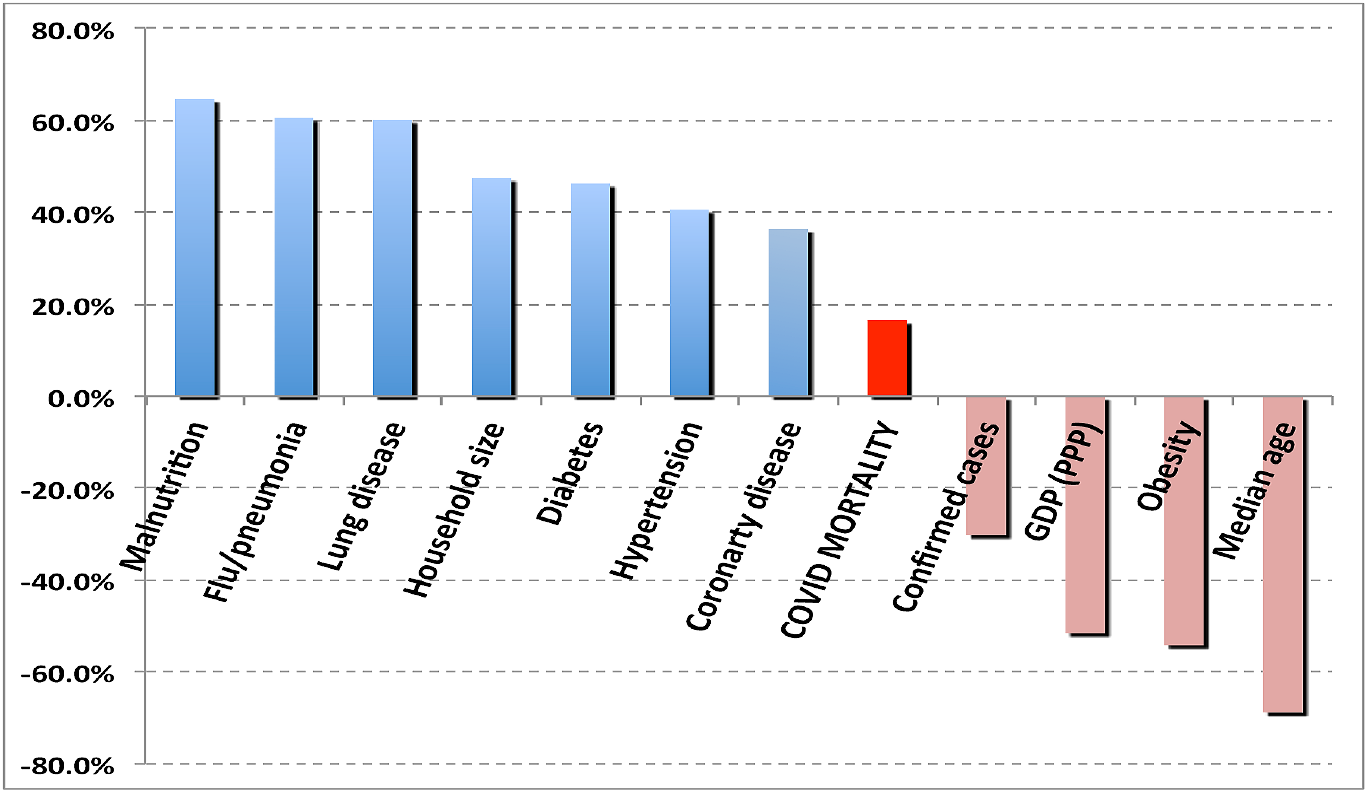
Cross-correlations of asthma with several disorders that might affect the outcome of COVID-19 cases.

Although obesity appears correlated with SARS-CoV-2 contagion, it appears uncorrelated with the outcome of COVID-19 infections contrary to the findings of reference [9]. Figure 13 shows that lack of such correlation does not appear with respect to influenza, malnutrition and asthma, although in those three cases the coefficient is negative. Understanding the correlations of obesity calls for a deeper look at the relation of obesity with the conditions that show the most influence. Already in the case of contagion, regional differences make for a substantial fraction of the apparent effect (Figure19).

**Figure 19.**
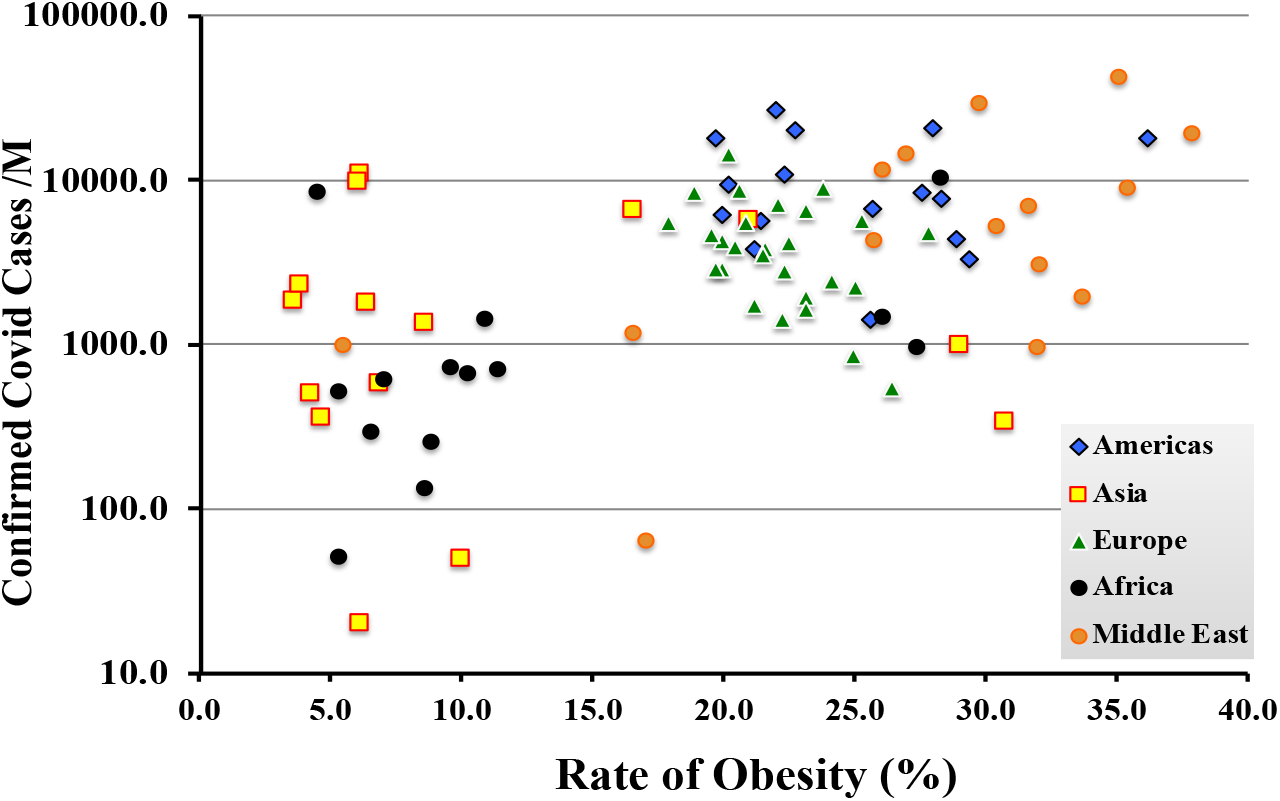
Is the correlation with obesity merely due to regional differences with other underlying influences?

The regional differences could be due to factors such as national median age (Figure 20) or it may be influenced by national wealth reckoned in terms of per capita GDP-PPP (Figure 23).

**Figure 20.**
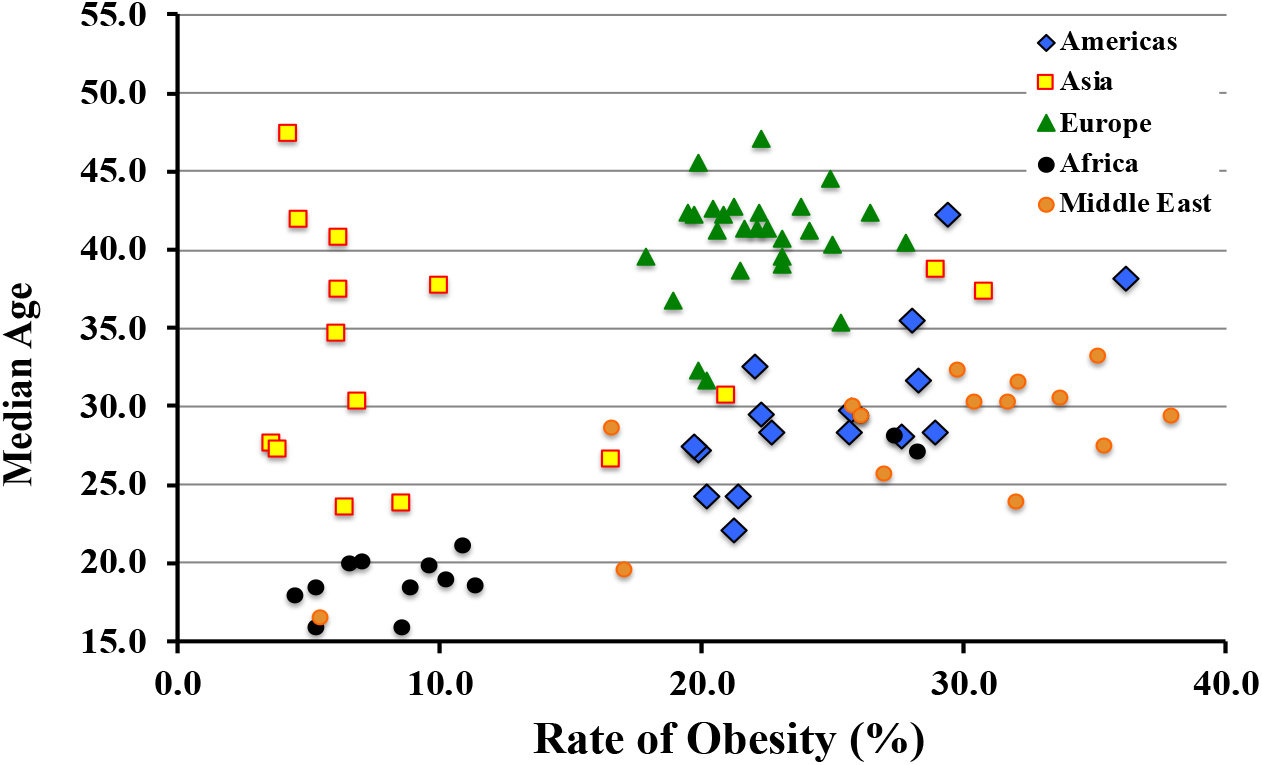
Correlation of obesity with national median age

**Figure 21.**
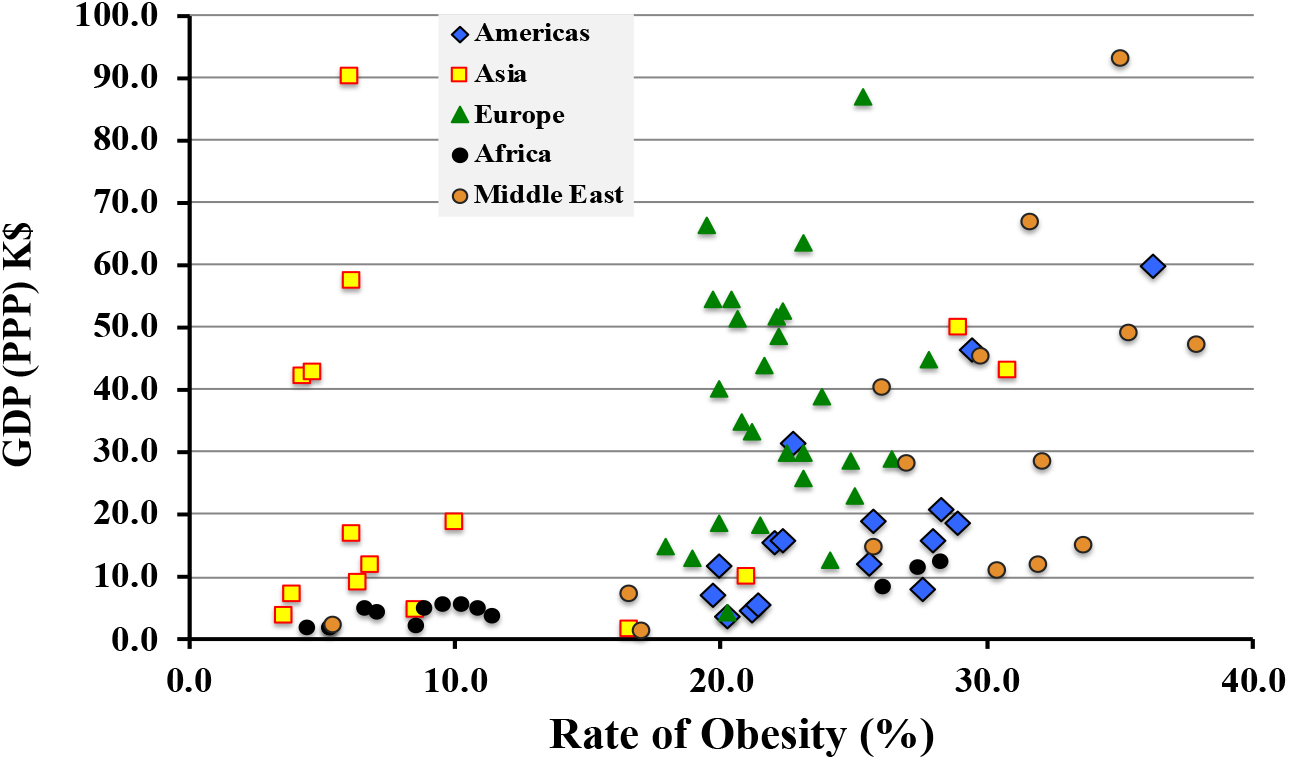
Or is wealth of the nation the driver? Correlation of obesity with GDP (PPP)

**Figure 22.**
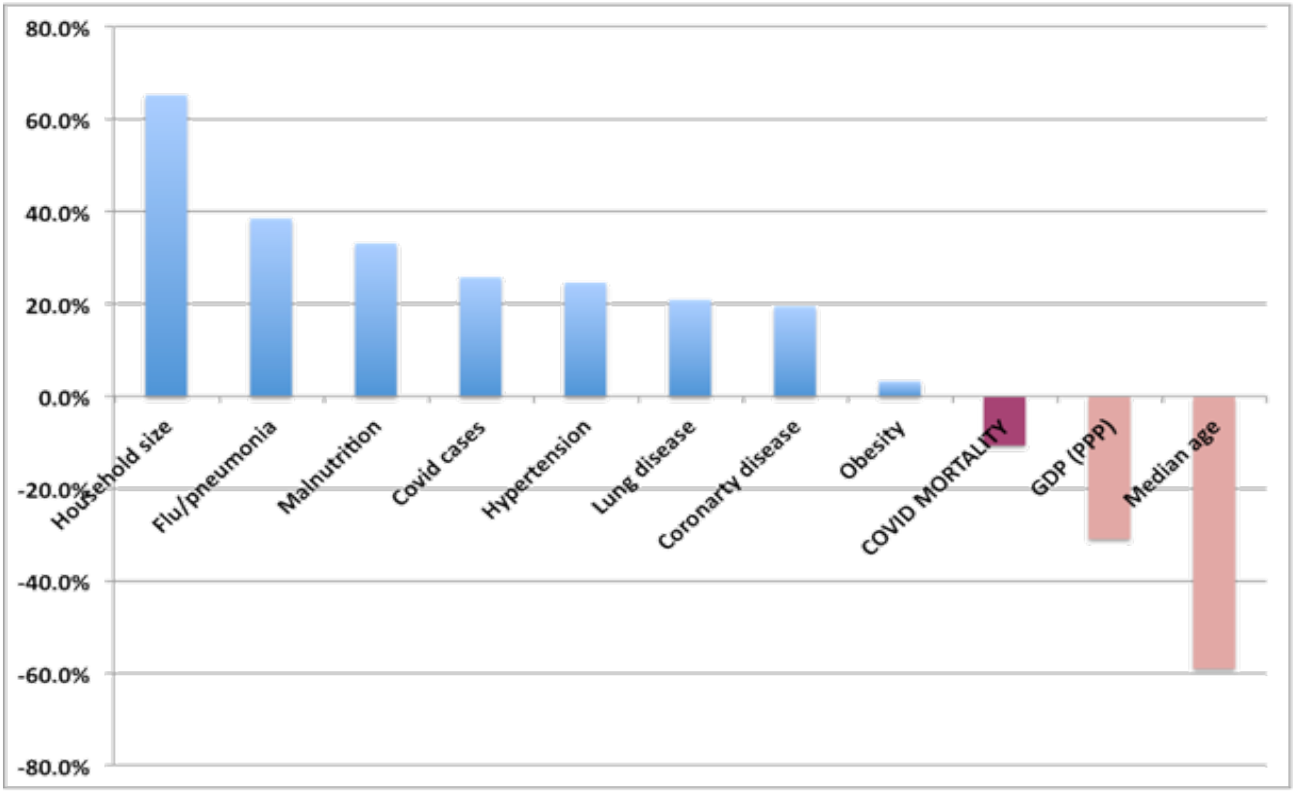
Correlations of diabetes mellitus with other medical and economic conditions. The correlation with COVID-19 mortality is the purple bar.

**Figure 23.**
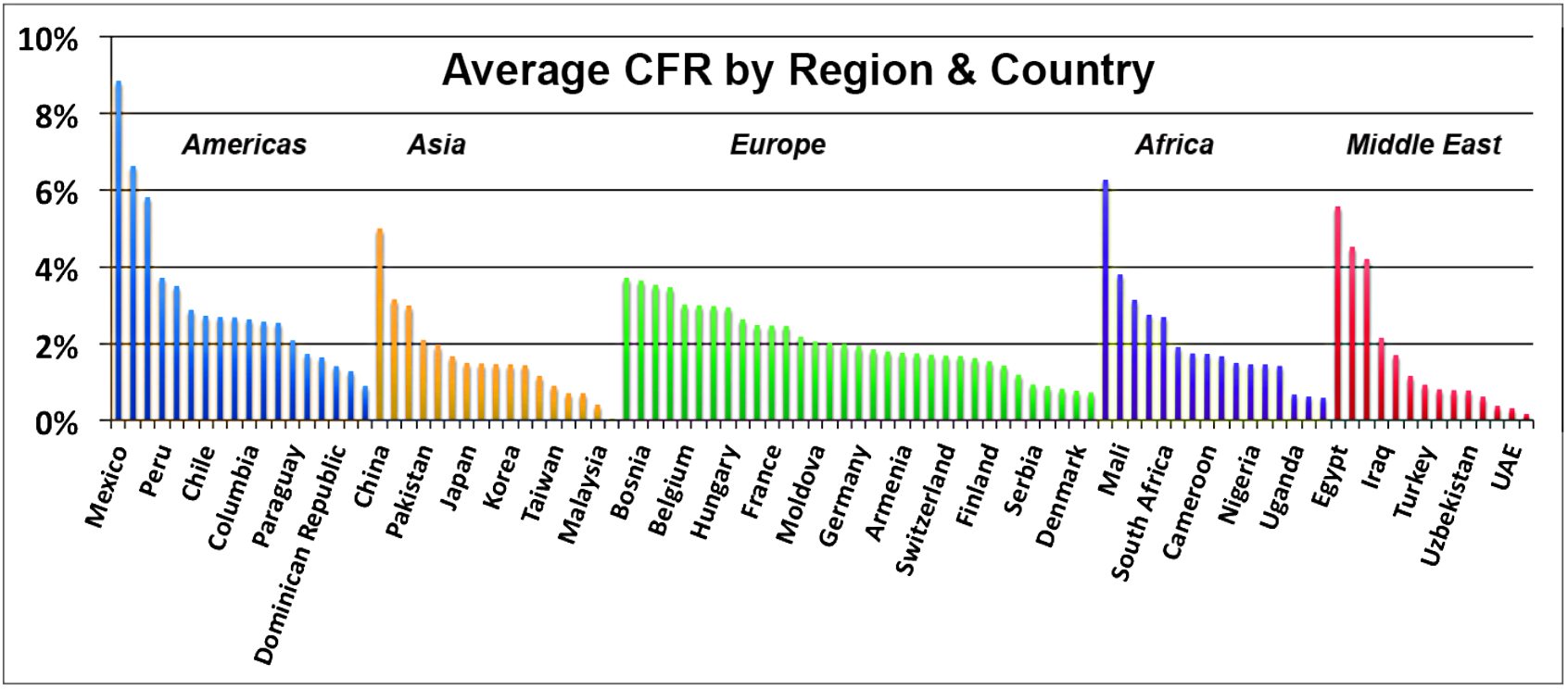
Average case fatality rate by country and region

Figures 19 through 21 do not uncover the means by which obesity may influence COVID-19 contagion, but they do illustrate why cross-correlations are important to examine.

As is the case with asthma, diabetes mellitus shows (Figure 22) significant correlations with several medical and economic conditions such as age, household size and mortality due to influenza/pneumonia. Once again no correlation with COVID-19 mortality is evident.

### 4.2 Regional and analysis

A key assumption of this study is the high degree of country dependence of the COVID-19 case fatality rate. Even though the CFR has fallen dramatically in many countries with rates originally greater than 10%, after nearly one year of pandemic, the disparity by country and by region remains large, ranging over an order of magnitude as illustrated in Figure 23.

The size of regional data sets is obviously much small than the aggregated world data. Consequently the uncertainties in computed correlations are higher. Nonetheless, examining the regional dependence of COVID-19 CFR on the most commonly cited co-morbidities is instructive. (Figure 24.)

**Figure 24.**
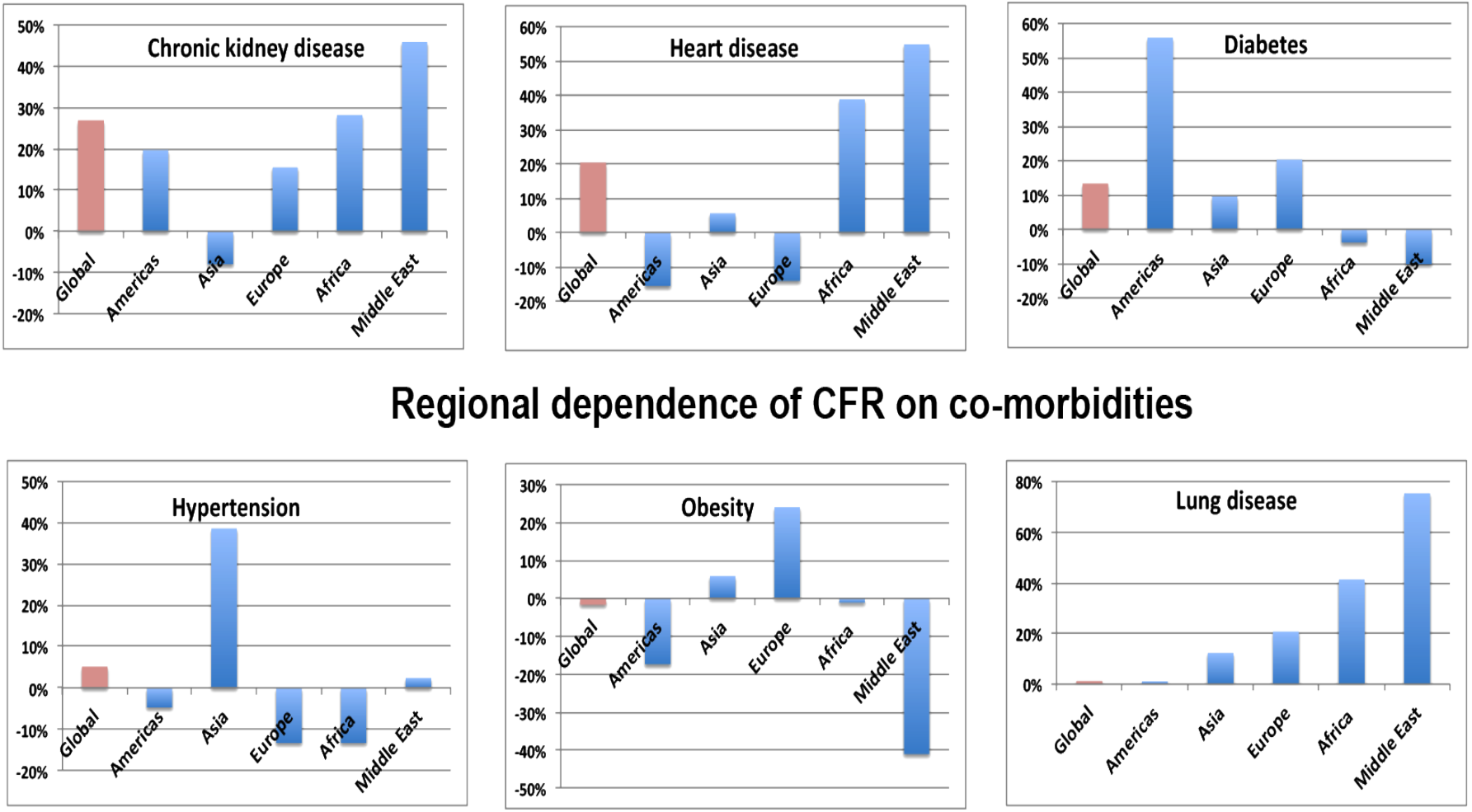
The regional dependence of commonly cited medical co-morbidities

### 5. Factors related to national economics and public health policies

The differences in the magnitude, outcomes, and characteristics of the waves of infections among sub-national regions with roughly equivalent medical factors indicates that economics and public health policies makes a significant difference in the severity of SARS-Cov-2 infections. This section examines dependencies on GDP-PPP, average household size, percentage of population living in slums, percentage of urban population, health expenditures per capita, and the WHO Universal Health Coverage (UHC) index.

Figure 4 has already shown an example of economic impact on medical outcomes; the per capita GDP (after correction for purchasing power) has a strong influence (−44.6%) on the rate of deaths due to malnutrition. That observation is hardly surprising. One may ask the same question with respect to mortality due to COVID-19 infections. Figure 25 shows essentially no correlation (−5.9%) of COVID-19 mortality with national wealth; the politics of poverty does not explain the observed national rates of COVID-19 mortality.

**Figure 25.**
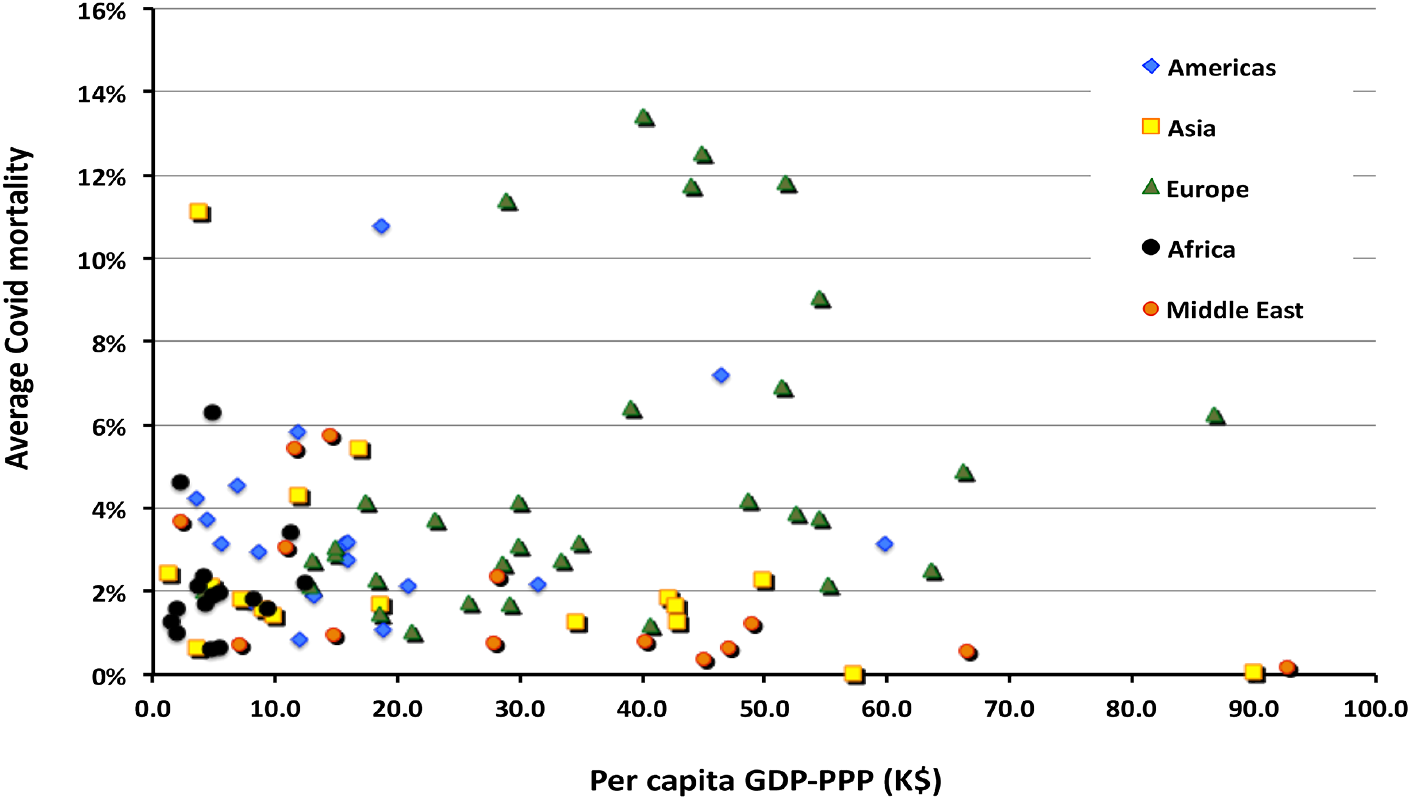
The distribution of COVID-19 mortality with national GDP. No correlation is observed in these data for any region.

The influence on contagion of SARS-Cov-2 over the entire data set is noticeable and positive (29.9%). As shown in Figure 26 that value is entirely driven by strong dependence of rising contagion with rising income in African countries. If one removes the African countries from the sample, the correlation disappears (2.8%).

**Figure 26.**
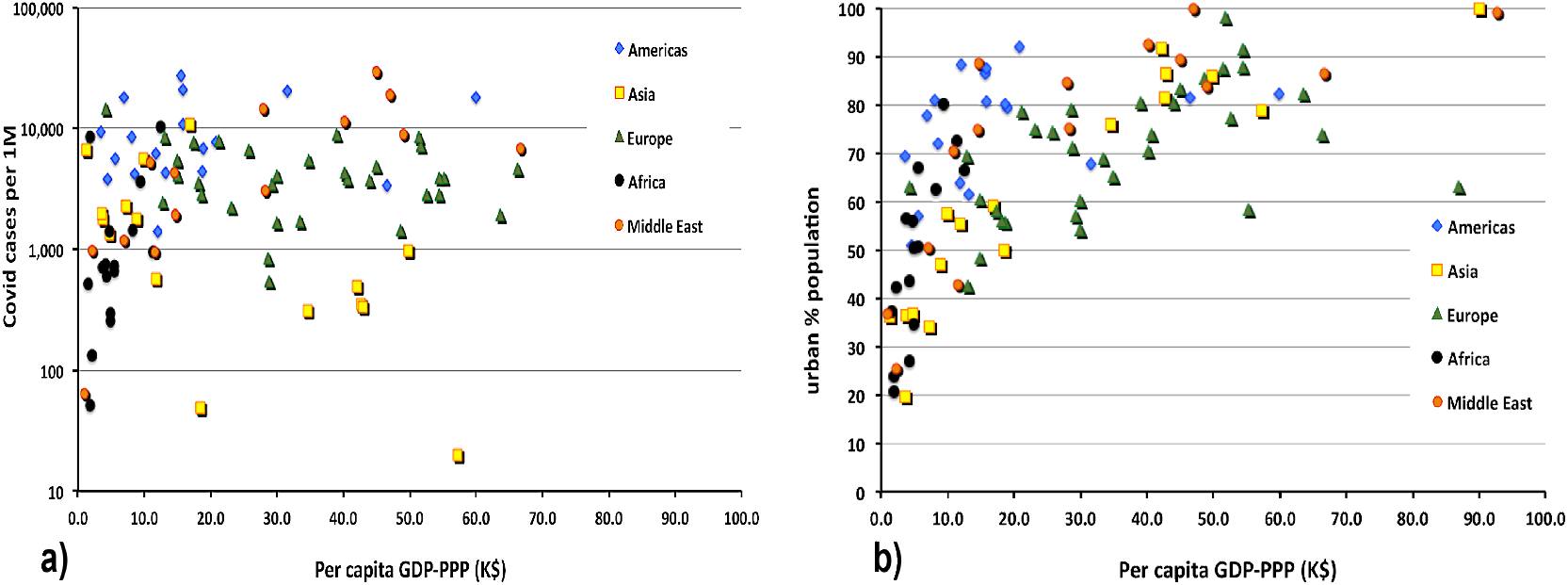
a) The distribution of COVID-19 cases with national GDP-PPP. b) The degree of urbanization with increasing GDP

The strange behavior in Africa in Figure 26a may be due to the increase in urbanization with increasing national wealth. One might further suspect that the increase in urbanization is also likely to increase the fraction of the national population living in slums. In fact, taken together Figure 26b and Figure 27 show that both of those suppositions are consistent with the data.

**Figure 27.**
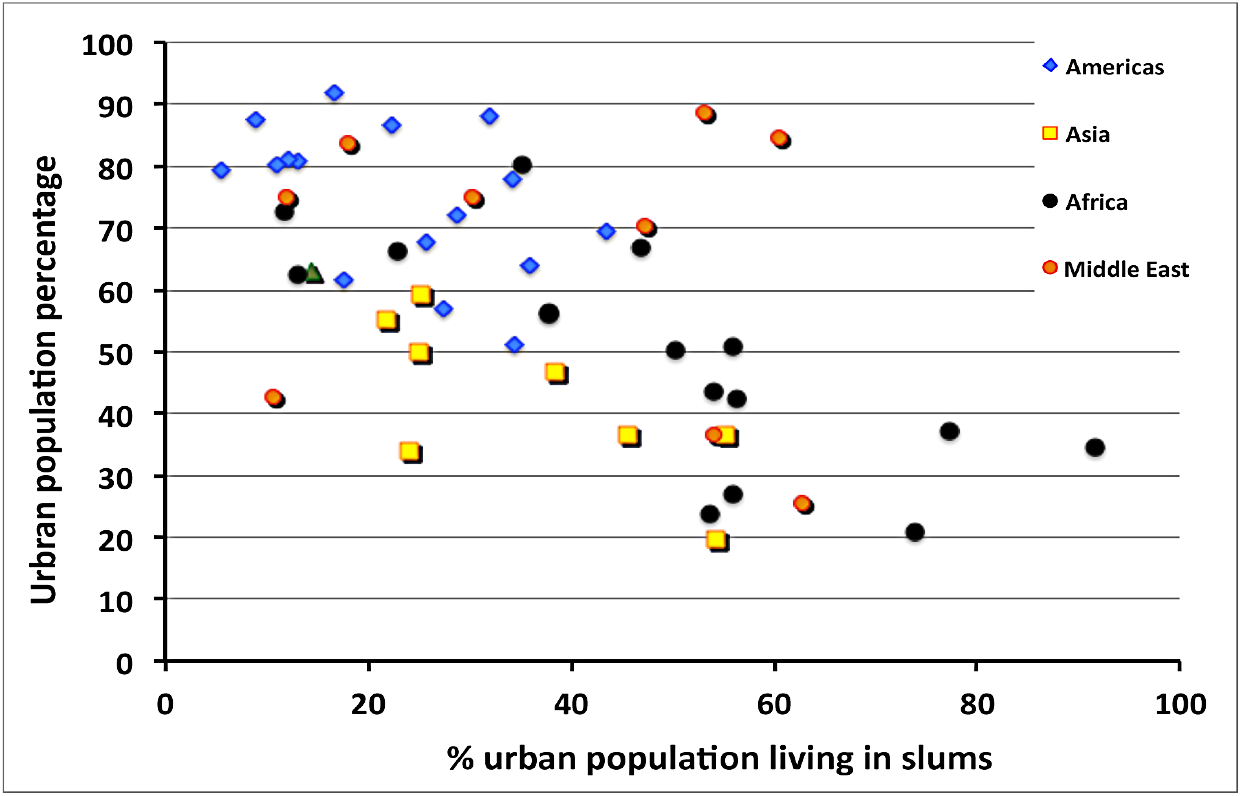
Correlation of the urban population percentage with the percentage of the urban population living in slums.

The correlation of economic and policy factors with contagion (measured in confirmed COVID-19 cases per 1 million of population and apparent COVID-19 mortality is presented in Table 5. As the mortality rate varies in time and seems to decline as the pandemic progresses (at least in the Northern Hemisphere) the mortality rate has been benchmarked on December 30, 2020.

**Table 5.** Correlations of economic and political factors with number of cases of COVID-19 infection (contagion) and apparent COVID-19 mortality.

| Factor | Correlation |  |
| --- | --- | --- |
|  | Contagion | Mortality |
| % in cities | 47.6% | -8.5% |
| Testing for COVID-19 | 43.8% | -9.9% |
| GDP-PPP | 32.0% | -5.9% |
| UHC INDEX | 30.3% | -2.0% |
| Health spending | 24.0% | 4.6% |
| Household size | 17.2% | 5.6% |
| Urban % in slums | -40.7% | 4.1% |

One may perform an examination by region for the impact of economic co-factors in the COVID-19 case fatality rate. That is shown in Figure 28.

**Figure 28.**
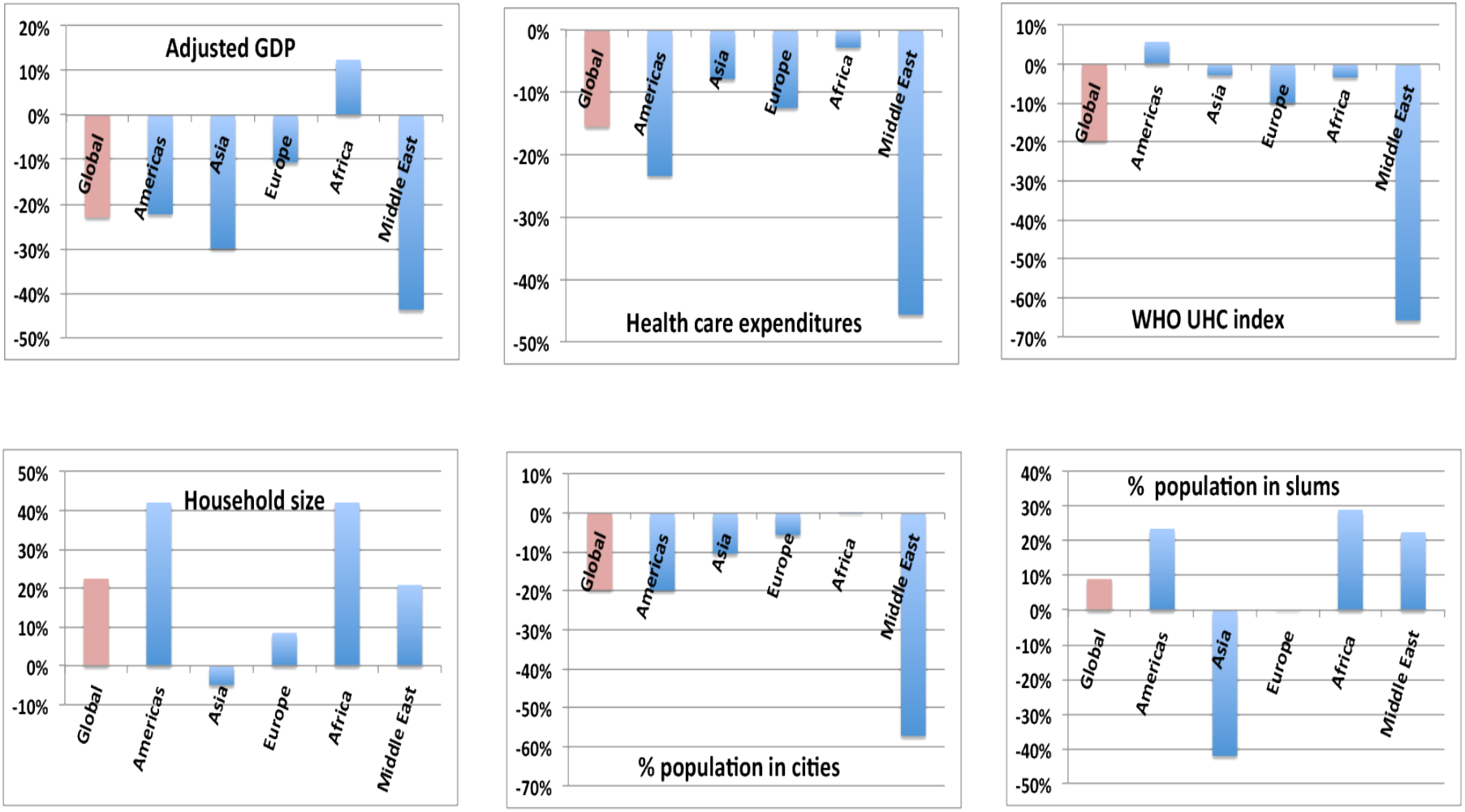
Impact of economic co-factors on CFR also vary strongly by region

The negative correlations with national wealth and with health care expenditures are to be expected. Nonetheless these effects are weaker in Africa than in other regions. More detailed discussion of these effects would require examination of underlying conditions on a country-by-country basis.

The surprising negative correlation in contagion with the percentage of the urban population living in slums is likely due to the trend in Africa that the smaller the fraction of the population living in cities, the more likely it is that they live in slums [World Bank data]. (Figure 27).

The correlation with respect to GDP is explained by the correlation of GDP with percentage of population over 65. The substantial correlation of contagion with testing results from the obvious fact that the more one looks, the more one sees. The correlation of contagion with percentage of urban population is due to the cross-correlation of GDP with percentage of urban population (64.8%) and the high cross-correlation of urban population with testing for COVID-19 (49.7%). The values for average health care expenditures and the UHC index of the WHO are similarly explained. The data that underlie the value of case fatality rate versus the percentage of the urban population that live in slums appears below in Figure 29.

**Figure 29.**
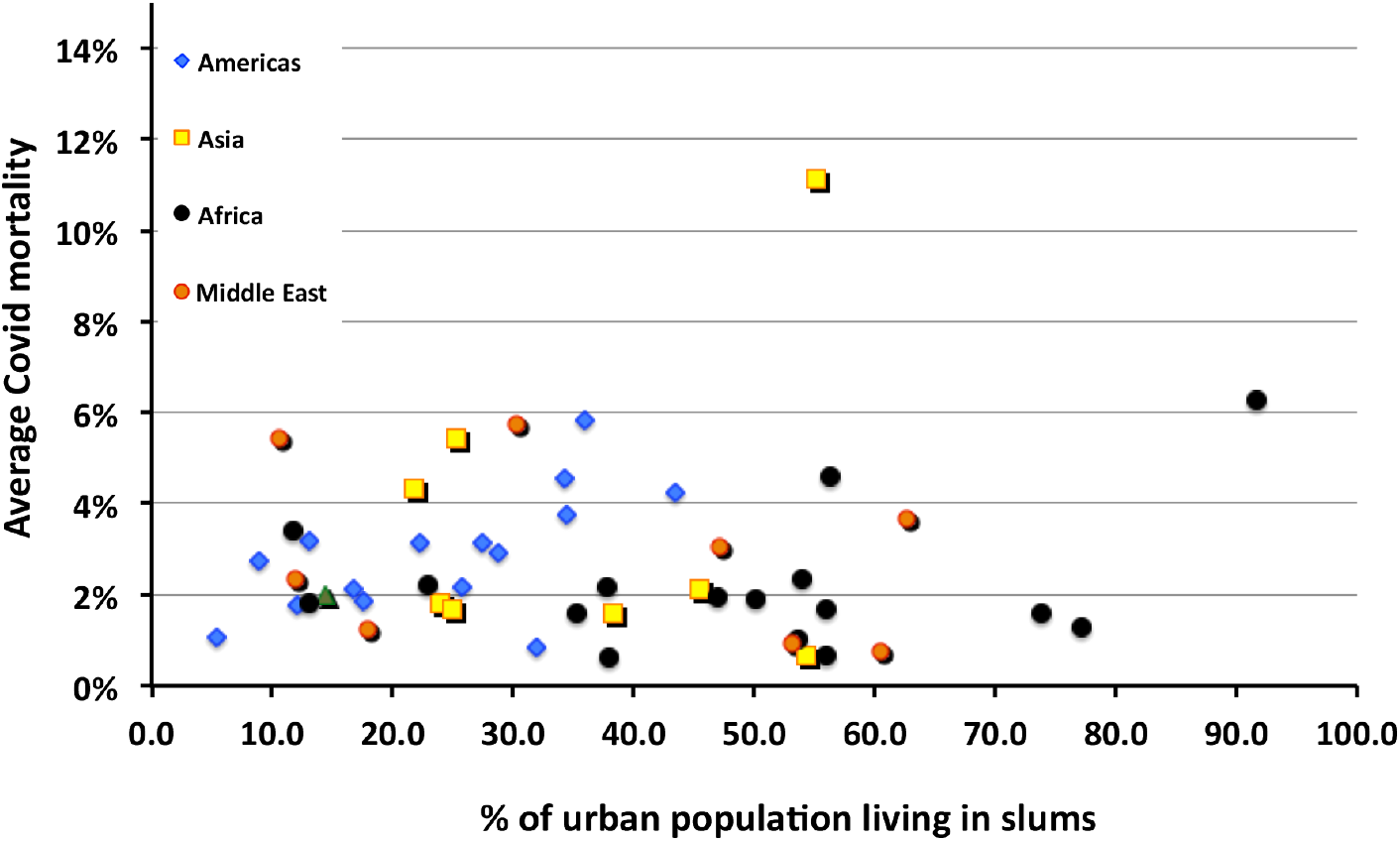
Correlation of COVID-19 mortality with the percentage of the urban population living in slums.

## 6. Summary and conclusions

This study covering statistics from countries with ∼70% of the world’s population confirms the early clinical observation that infection by the SARS-Cov-2 virus presents an increased risk to persons over the age of 65. However, it does not support the suggestions presented by government agencies early in the pandemic that the risks are much greater for persons with certain common, potential co-morbidities. Many of the early deaths of elderly patients early in the course of the pandemic took place in circumstances that likely promoted rather than impeded the spread of the virus among person who were already in a generally poor state of health, likely accompanied by compromised immune function.

A plausible explanation is provided in references [4] and also by Koff and Williams in [8]; namely the virulence of COVID-19 in the elderly is strongly driven by the decrease in adaptive and innate immune responses with aging. Koff and Williams go on to recommend more longitudinal studies in aging populations including assessing the potential of a decrease in the efficacy of vaccines as being “critical to the future of global health.”

A commonly heard claim by many persons who object to strict measures to prevent the spread of the SARS-CoV-2 virus has been that the resulting disease is similar to influenza, is only slightly more lethal, and should be treated in the same manner as influenza as a matter of public policy. The comparison of the severity of medical outcomes of COVID-19 with those caused influenza strains and their resulting pneumonias displays dramatic differences. Promulgating the idea that COVID-19 a “flu-like disease” spreads gross misinformation to the detriment of the public health worldwide.

One may ask what governmental actions can reduce the seriousness of infections by SARS-CoV-2. Comparing the cases of Germany and Italy may be instructive in this regard. Both countries have similar numbers of confirmed cases of COVID-19; yet the death rate in Italy is roughly triple that in Germany. Germany put in place and extensive number of triage and early treatment centers outside of hospitals; Germany also moved quickly to secure adequate supplies of personal protective equipment. Infected patients were identified early in the course of the disease and were treated in a manner that did not overwhelm the central intensive care facilities in hospitals as happened in the Italian region of Lombardia.

Perhaps a similar lesson comes from comparing the experience in the United States in California and New York. The early lockdown in California more than doubled the first duration of the first wave of infections as compared with New York leading to 60% more cases in California yet half the death rate of New York in which medical resources were badly stressed.

Presently authoritative data on a worldwide, country-to-country basis are not publicly available to evaluate the effectiveness of either prevention and/or treatment modalities. Also unavailable over the full range of those countries included in this analysis are the complete statistics related to COVID-19 disaggregated with respect to sex. When and if such data become available, expanding the analysis with respect to sex-based differences in testing, contagion, and mortality would prove useful.

The roll out of large-scale vaccination programs during a time when the vaccines are in short supply has necessitated the schemes for prioritizing recipients. If probability of sever illness is a primary consideration, then the early guess about the risks connected with potential co-morbidities should be replaced with data such as presented here along with detailed clinical evaluations accumulated throughout 2020.

A last word of caution: the data used in this study have been accumulated before the UK (B117) and South African (V2) strains of SARS-CoV-2 began to propagate. Already early in 2021, some evidence is suggesting that these new strains maybe be somewhat more virulent. Figure 30 plots the case fatality rate averaged from the time of the first clusters of COVID-19 for several countries.

**Figure 30.**
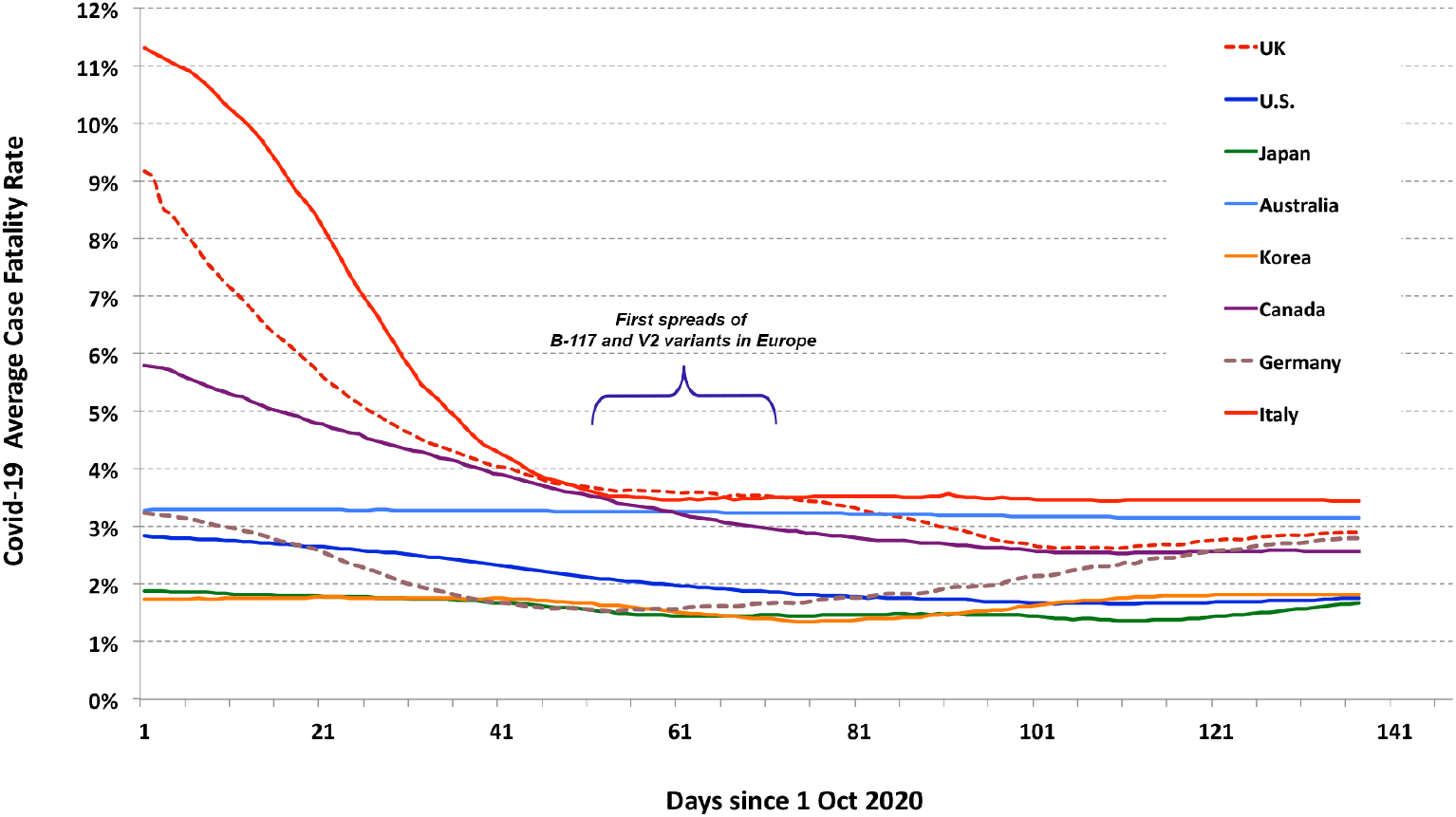
The case fatality rates for several countries averaged from the time of the appearance of first clusters of cases.

One sees a troubling trend for the average CFR to begin to increase. The period of the first spread of viral variants is indicated as a time marker. It is too soon to judge whether the upturn whether is a reflection of increased virulence in variants of SARS-CoV-2, whether it is an indication of increased susceptibility and physical and psychological stress on so-called “essential workers,” or whether it is a result of some form of COVID weariness among large portions of national populations. Difference in virulence of the several variant strains now circulating will complicate the interpretation of national data collected in 2021,

## Data Availability

All data in this article are in the public domain

https://www.worldometers.info/coronavirus/#countries

https://www.worldlifeexpectancy.com/

## Acknowledgements

The author acknowledges his colleagues in the World Federation of Scientists for their encouragement to continue, expand, and report this research. The author’s work is completely self-supported without any outside funding.

---

**Table A.1.**
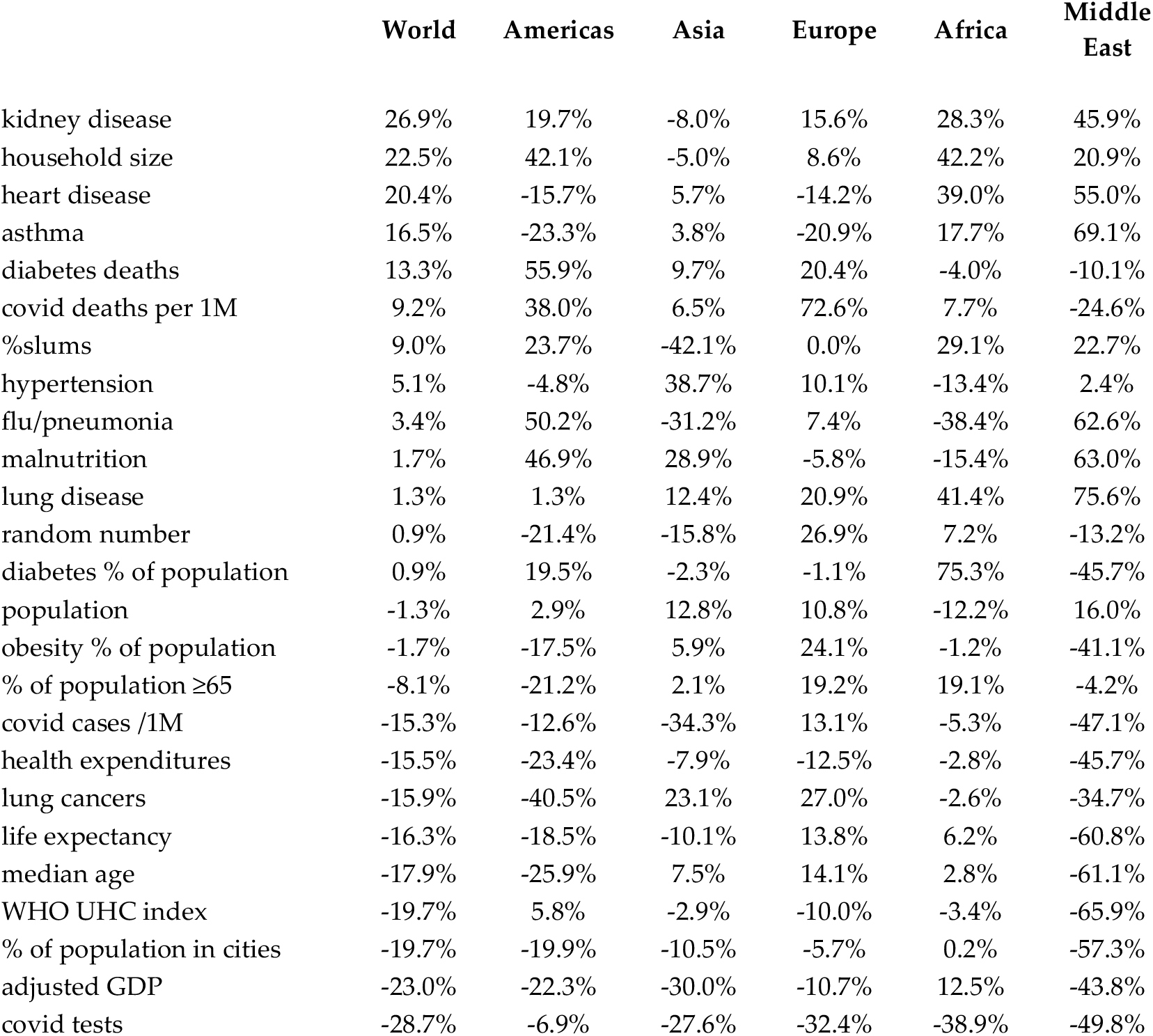
Regional variation of COVID-19 CFR correlations

|  | **World** | **Americas** | **Asia** | **Europe** | **Africa** | **Middle East** |
| --- | --- | --- | --- | --- | --- | --- |
| kidney disease | 26.9% | 19.7% | -8.0% | 15.6% | 28.3% | 45.9% |
| household size | 22.5% | 42.1% | -5.0% | 8.6% | 42.2% | 20.9% |
| heart disease | 20.4% | -15.7% | 5.7% | -14.2% | 39.0% | 55.0% |
| asthma | 16.5% | -23.3% | 3.8% | -20.9% | 17.7% | 69.1% |
| diabetes deaths | 13.3% | 55.9% | 9.7% | 20.4% | -4.0% | -10.1% |
| covid deaths per 1M | 9.2% | 38.0% | 6.5% | 72.6% | 7.7% | -24.6% |
| %slums | 9.0% | 23.7% | -42.1% | 0.0% | 29.1% | 22.7% |
| hypertension | 5.1% | -4.8% | 38.7% | 10.1% | -13.4% | 2.4% |
| flu/pneumonia | 3.4% | 50.2% | -31.2% | 7.4% | -38.4% | 62.6% |
| malnutrition | 1.7% | 46.9% | 28.9% | -5.8% | -15.4% | 63.0% |
| lung disease | 1.3% | 1.3% | 12.4% | 20.9% | 41.4% | 75.6% |
| random number | 0.9% | -21.4% | -15.8% | 26.9% | 7.2% | -13.2% |
| diabetes % of population | 0.9% | 19.5% | -2.3% | -1.1% | 75.3% | -45.7% |
| population | -1.3% | 2.9% | 12.8% | 10.8% | -12.2% | 16.0% |
| obesity % of population | -1.7% | -17.5% | 5.9% | 24.1% | -1.2% | -41.1% |
| % of population ≥65 | -8.1% | -21.2% | 2.1% | 19.2% | 19.1% | -4.2% |
| covid cases /1M | -15.3% | -12.6% | -34.3% | 13.1% | -5.3% | -47.1% |
| health expenditures | -15.5% | -23.4% | -7.9% | -12.5% | -2.8% | -45.7% |
| lung cancers | -15.9% | -40.5% | 23.1% | 27.0% | -2.6% | -34.7% |
| life expectancy | -16.3% | -18.5% | -10.1% | 13.8% | 6.2% | -60.8% |
| median age | -17.9% | -25.9% | 7.5% | 14.1% | 2.8% | -61.1% |
| WHO UHC index | -19.7% | 5.8% | -2.9% | -10.0% | -3.4% | -65.9% |
| % of population in cities | -19.7% | -19.9% | -10.5% | -5.7% | 0.2% | -57.3% |
| adjusted GDP | -23.0% | -22.3% | -30.0% | -10.7% | 12.5% | -43.8% |
| covid tests | -28.7% | -6.9% | -27.6% | -32.4% | -38.9% | -49.8% |

## Footnotes

1 World Health Rankings, https://www.worldlifeexpectancy.com/world-health-rankings

2 Worldometer, https://www.worldometers.info/

3 Trading Economics, https://tradingeconomics.com/

4 The GDP-PPP has a several percent systematic uncertainty that depends on the economic model used in making the PPP corrections.

## References

[1] “People with Certain Medical Conditions,” https://www.cdc.gov/coronavirus/2019-ncov/need-extra-precautions/people-with-medical-conditions.html, August 14, 2020

[2] H. Li, S. Wang, F., Zhong, W. Bao, Y. Li, L. Liu, H. Wang and Y. He, “Age-Dependent Risks of Incidence and Mortality of COVID-19 in Hubei Province and Other Parts of China, Front. Med. (Lausanne) v.7, p.190; 2020

[3] U.S. CDC, “COVID-19 Hospitalization and Death by Age,” https://www.cdc.gov/coronavirus/2019-ncov/covid-data/investigations-discovery/hospitalization-death-by-age.html, August 18, 2020, 3.

[4] E. Montecino-Rodriguez, B. Berent-Maoz, and K. Dorshkind, Causes, consequences, and reversal of immune system aging,” J Clin Invest. 2013 Mar 1; 123(3): 958–965.

[5] M. Kenji, K. Katsushi, Z. Alexander, and C. Gerardo. “Estimating the asymptomatic proportion of coronavirus disease 2019 (COVID-19) cases on board the Diamond Princess cruise ship, Yokohama, Japan,” 2020. Euro Surveill. 2020;25 (10):pii=2000180. https://doi.org/10.2807/1560-7917.

[6] The report of the UK government website notes several caveats: 1) The figures include deaths of non-residents of the UK; 2) they are based on the date that a death was registered rather than occurred; 3) they are provisional and use the tenth edition of the International Classification of Diseases, (ICD-10) for definitions for the coronavirus (COVID-19). https://www.ons.gov.uk/peoplepopulationandcommunity/healthandsocialcare/conditionsanddiseases/articles/coronaviruscovid19roundup/2020-03-26. See also J. P.A. Ioannidis, C. Axfors, D. G. Contopoulos-Ioannidis, “Population-level COVID-19 mortality risk for non-elderly individuals overall and for non-elderly individuals without underlying diseases in pandemic epicenters,” Environmental Research, ISSN: 0013-9351, Vol: 188, Page: 109890

[7] S. de Lusignan, J. Dorward, Ana Correa, et al., “Risk factors for SARS-CoV-2 among patients in the Oxford Royal College of General Practitioners Research and Surveillance Centre primary care network: a cross-sectional study,” Lancet Infect Dis., 2020 Sep; 20(9): 1034–1042. Published online 2020 May 15. doi: 10.1016/S1473-3099(20)30371-6

[8] W.C. Koff and M. A. Williams, “COVID-19 and Immunity in Aging Populations— A New Research Agenda,” N Engl J Med 383;9 (Aug. 27, 2020)

[9] A. Tamara and D. L. Tahapary, “Obesity as a predictor for a poor prognosis of COVID-19: A systematic review,” Diabetes & Metabolic Syndrome: Clinical Research & Reviews, V.4, Issue 4, July–August 2020, Pages 655–659

